# The impact of the COVID-19 pandemic on hospital services for patients with cardiac diseases: a scoping review

**DOI:** 10.1101/2021.12.01.21267100

**Authors:** Mats de Lange, Ana Sofia Carvalho, Óscar Brito Fernandes, Hester Lingsma, Niek Klazinga, Dionne Kringos

## Abstract

**Background:** The repercussions of the COVID-19 pandemic concern care in many clinical areas, including cardiology. We aim to assess the impact of the COVID-19 pandemic on hospital care for cardiac patients.

**Methods:** Scoping review. Performance indicators were extracted and collated to inform on changes in the use of health services and care provided during January - June 2020.

**Results:** Database searches yielded 6277 articles, of which 838 articles met the inclusion criteria during initial screening. After full-text screening, 94 articles were considered for data extraction. In total, 1637 indicators were retrieved, showing large variation in the indicators and their definitions. Most of the indicators that provided information on changes in number of admissions (n=118, 88%) signalled a decrease in admissions; 88% (n=15) of the indicators showed patients’ delayed presentation and 40% (n=54) showed patients in a worse clinical condition. A reduction in diagnostic and treatment procedures was signalled by 95% (n=18) and 81% (n=64) of the indicators reporting on cardiac procedures, respectively. Length of stay decreased in 58% (n=21) of the indicators and acute coronary syndromes treatment times increased in 61% (n=65) of the indicators. Outpatient activity decreased in 94% (n=17) of the indicators related with outpatient care, whereas telehealth utilization increased in 100% (n=6). Outcomes worsened in 40% (n=35) of the indicators, and mortality rates increased in 52% (n=31).

**Conclusion:** All phases of the hospital cardiac care pathway were affected. This information could support the planning of care during the ongoing pandemic and in future events.

## Introduction

As of 12 January 2022, the virus causing COVID-19 disease, SARS-CoV-2, has infected 313 million people globally and caused 5.5 million deaths (1-3). The use of health resources for non-COVID care was minimized, elective procedures and appointments were postponed since the beginning of 2020 (4,5). The impact of COVID-19 on health care services for patients with non-communicable diseases (NCDs) seems to be severe and involving multiple clinical areas (6-12).

According to the WHO, cardiovascular diseases, which include cardiac diseases, are the leading cause of death globally (13). Before the pandemic, a decelerating pace in the improvement of cardiovascular disease mortality was already identified as a major contributor to the slowdown of life expectancy gains in several Organisation for Economic Co-operation and Development (OECD) member countries (14). The continuity of care for patients with cardiac diseases during crisis is a major concern among healthcare providers (15,16), who seek to strengthen referrals and care pathways, and establishing and maintaining novel models of care. Given the burden of cardiac diseases in healthcare systems globally (13,14), attention for the impact of the COVID-19 pandemic on cardiology patients is justified. Previous studies noticed a decrease in patients presenting with (acute) cardiac conditions and mentioned delays in those patients who eventually did present for hospital care (17-19), which could lead to worse clinical outcomes (20).

In a relatively short period, large amounts of scientific evidence have become available on the impact of the pandemic on patients with cardiac diseases in varying countries. This literature is scattered, which makes it difficult to draw conclusions regarding the impact on cardiac care delivery and the use of data to steer health care services delivery improvements during the current pandemic and future public health crises. Hence, reviewing the existing literature that quantifies the magnitude of this impact during the first half of 2020 helps to systematize, synthesize, and consolidate the evidence, and to formulate recommendations for policy and practice.

This scoping review is part of a larger project that focuses on performance indicators used for a range of clinical areas during the COVID-19 pandemic. In this study, we aim to assess the impact of the pandemic on hospital care for cardiac patients in OECD countries, focusing on the different phases of the hospital cardiac care pathway (admission, diagnosis, treatment, outpatient care, outcomes).

## Methods

We pursued a scoping review methodology, considering the extensive number of scientific articles, their heterogeneous character, and the lack of a clear and structured overview of the large sums of literature that have become available on the impact of the pandemic on patient with cardiac conditions. A scoping review can be used to examine emerging evidence and give an overview of the literature and studies available on a certain theme (21). We followed the methodological framework developed by Arksey and O’Malley(22), further developed by Levac et al (23), and the PRISMA extension for scoping reviews to report our methodology (24) (Supplementary material S1).

### Search strategy

The MEDLINE and Embase databases were selected to search for this scoping review, as we considered them to sufficiently cover the literature related with delivery of health care services. Pilot searches were conducted to identify a list of suitable search terms. A medical research librarian was consulted to improve the search strategy and adapt it to both databases. The final search strategy included the following key terms and synonyms: COVID-19, pandemic, non-communicable disease, chronic disease, performance indicator, healthcare quality, healthcare utilization, healthcare delivery and other closely related terms. The full search strategy for Embase and MEDLINE can be found in Supplementary material S2. The comprehensive search was conducted by the research librarian on 17-03-2021. No limitations were set regarding language or year of publication. Duplicates were removed using EndNote software. Additional articles of relevance were added by hand-searching the reference lists of the included studies.

### Study selection

The following inclusion criteria were set: 1) studies using empirical data on the use of health services; 2) studies had to describe health outcomes and/or performance indicators during the COVID-19 pandemic; 3) studies that are presented as original journal articles using quantitative or qualitative methods, such as cohort studies, case-control, cross-sectional designs, case reports, systematic reviews, surveys, and meta-analyses. The following exclusion criteria were set: 1) non-primary studies (such as editorials and commentaries); 2) prediction models; 3) clinical case reports; and 4) diseases management or health services organization guidelines; 5) studies about the impact on healthcare workers, patients diagnosed with COVID-19, children, or pregnant women; 6) studies primarily performed in non-OECD countries; 7) articles from which only an abstract was available.

### Methods of selection

An initial screening of the retrieved studies based on title and abstract was performed independently by two researchers (ASC, OBF) using Rayyan (https://www.rayyan.ai/). Studies considered relevant after this phase were exported to a spreadsheet to support full-text screening, which was performed independently by three researchers (MdL, ASC, OBF). For this study only articles on cardiac diseases were analysed. The reason for exclusion of articles was recorded at this point. In case of doubt, the other co-authors were consulted, and a decision was made.

### Data extraction and charting

Data extracted from the included articles were collated in a spreadsheet informed by a pilot on 15 studies (Supplementary material S3). Data extraction was performed independently by three researchers (MdL, ASC, OBF). Extracted data included information on generic and methodological aspects of the article (e.g., authors, title, setting), and information about the indicators reported (e.g., indicator title, and data inclusion/exclusion considerations). For every indicator, we identified the trend reported in the articles (increase/decrease/stable).

### Synthesis of the results

Indicators were grouped and categorised according to the different phases of the hospital cardiac care pathway by MdL, followed by a review conducted by ASC (Supplementary material S4). The percentages of indicators showing a decreasing, increasing or stable trend were computed for each category. Results are presented in line with the hospital cardiac care pathway.

## Results

Database searches yielded 6277 articles. Of these, 838 articles focusing on non-communicable diseases met the inclusion criteria. After full-text screening of 117 articles focused on cardiac hospital care, a total of 94 articles were included in this review (Fig. 1). Twenty-three full-text studies were excluded.

**Figure 1.**
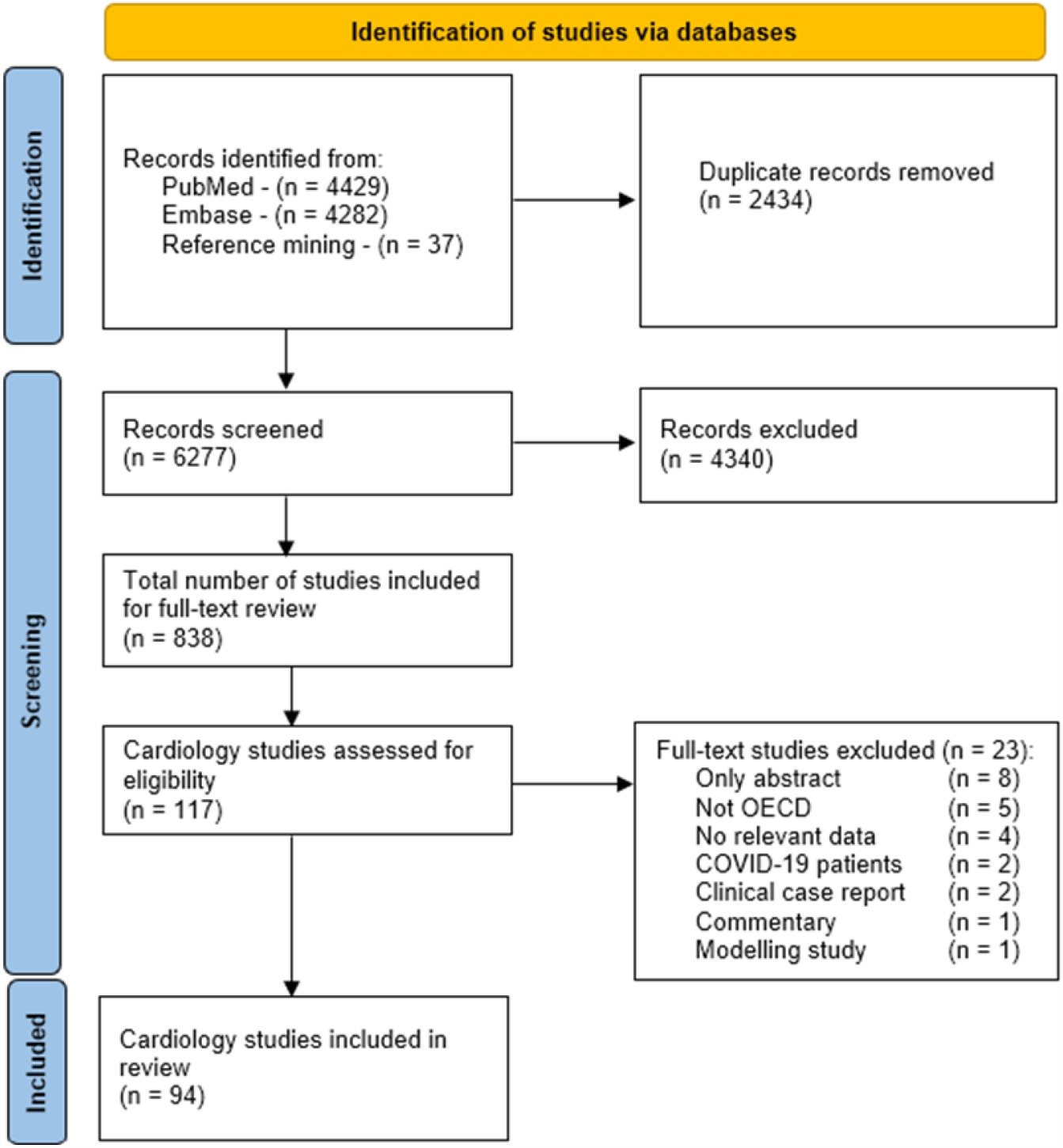
PRISMA flow diagram of the literature search. Abbreviations: OECD - Organisation for Economic Co-operation and Development.

### General characteristics of the included articles

The included articles reported on 109 different countries (Fig. 2) Eighty-six articles provided information on one country only. Eight articles involved multiple countries, of which seven also included non-OECD countries. Most articles reported on Italy (n = 20, 21%), followed by the United Kingdom (n = 17, 18%), and Germany (n = 14, 15%).

**Figure 2.**
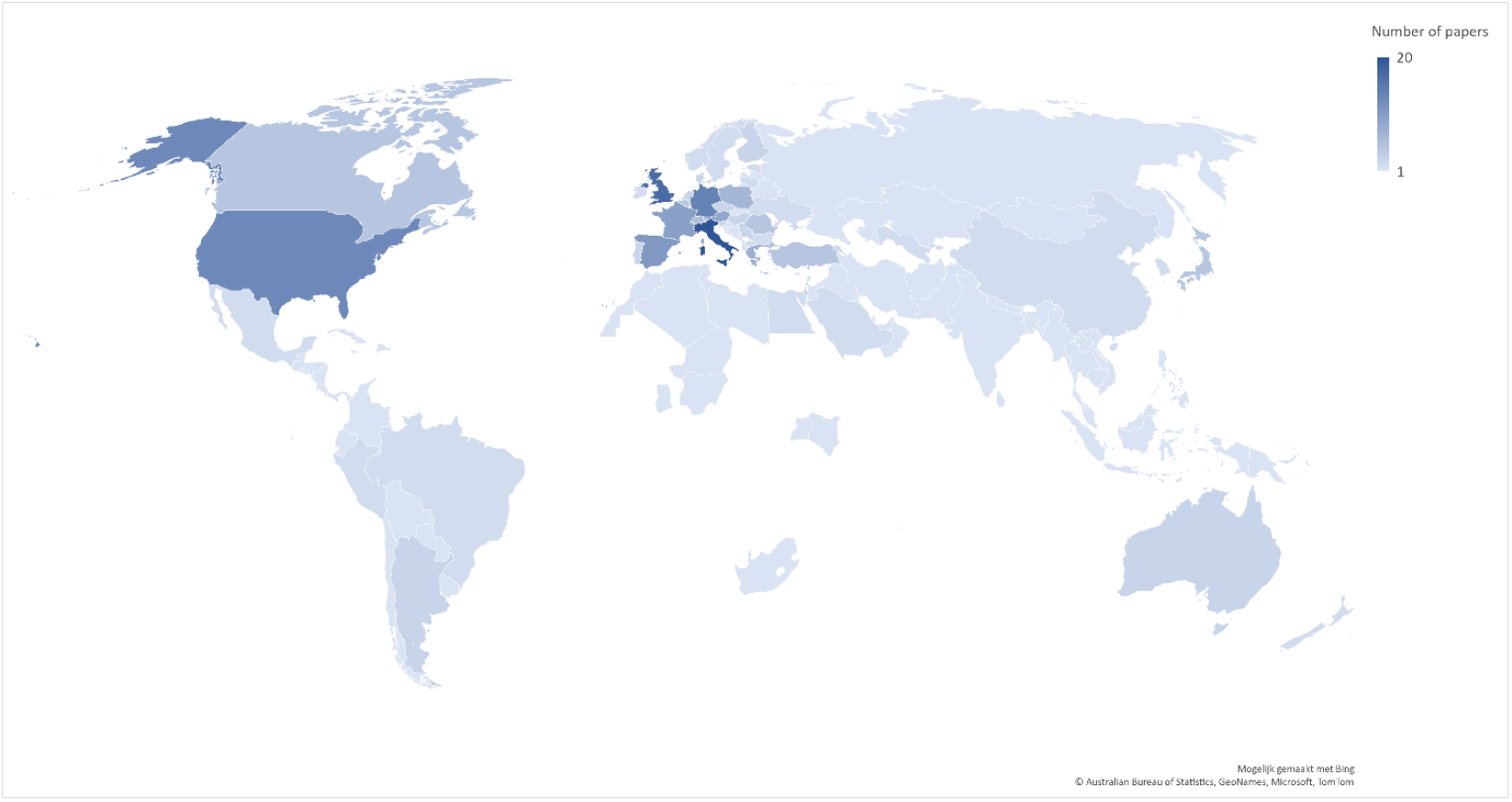
Countries reported on in the included articles (including articles reporting on multiple countries), colour-graded according to number of included papers (n=109)

Most of the studies used a retrospective cohort design (n = 66, 70%), other studies used a prospective cohort (n = 13, 14%), or survey design (n = 12, 13%). The included studies used three different time periods when comparing pandemic versus pre-pandemic indicators. Most studies compared a COVID-19 affected period in 2020 to the same period in the previous year (n = 54, 56%). Other studies (n = 34, 36%) compared the COVID-19 affected period to a period immediately before. Lastly, a COVID-19 affected period was compared to the average of the same period in several previous years (n = 27, 29%). The general characteristics for each article can be found in detail in Supplementary material S5.

### Impact of COVID-19 on the hospital cardiac care pathway

The grouped and categorised indicators were collated according to the different phases of the hospital cardiac care pathway (Fig. 3), to visualise their combined trends.

**Figure 3.**
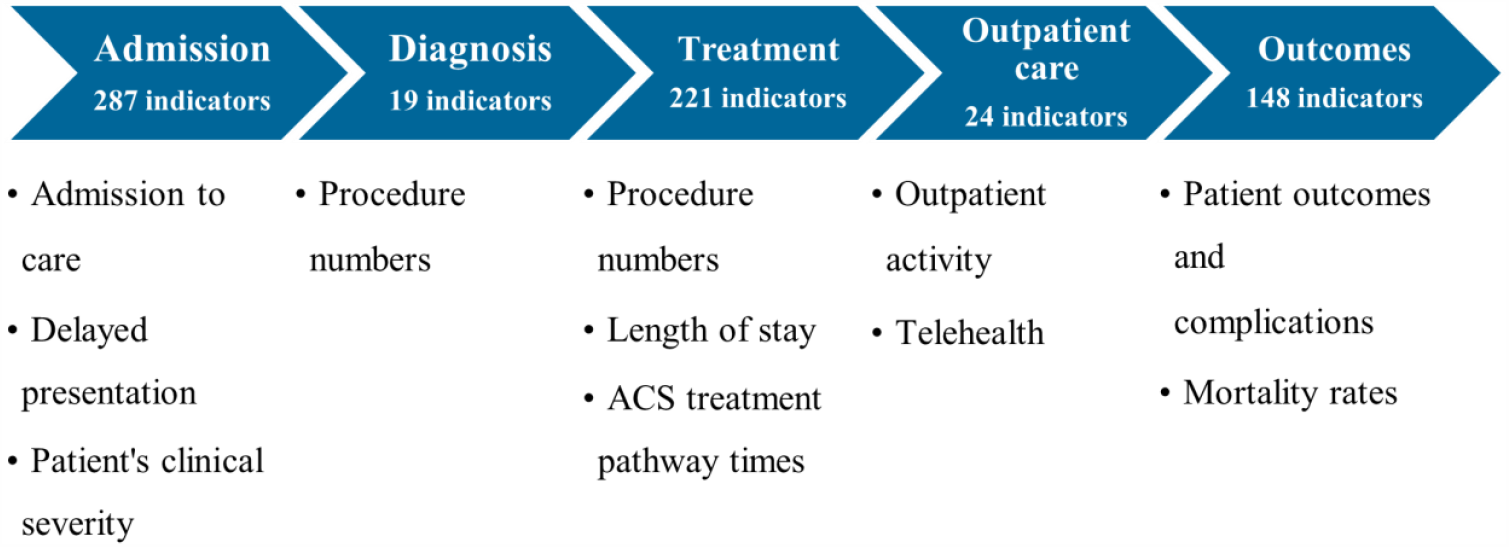
Categorisation of indicators according to different phases of the hospital cardiac care pathway. Abbreviations: ACS - Acute Coronary Syndrome

## Admission

A total of 287 indicators regarding the ‘admission’ phase of the hospital cardiac care pathway were identified. These indicators were grouped into three separate categories: ‘admission to care’, ‘delayed presentation’, and ‘patients’ clinical severity at presentation’.

### Admission to care

Regarding numbers of admissions for cardiac diseases to hospital services, 134 indicators were identified from 49 articles. Most of the indicators concerned patients with acute coronary syndrome (ACS), heart failure, or cardiac arrythmia. Of these indicators, 118 (88%) reported a decrease in number of admissions, compared to a non-COVID-19 period, 13 indicators (9.7%) reported a stable number of admissions, whereas 3 indicators (2.2%) reported an increase in admission numbers.

### Delayed presentation

Regarding the patients’ timing of presentation to hospital services, 17 indicators were identified from 12 articles. All the indicators in this category reported on patients with ACS. Most indicators defined delayed presentation as more than twelve hours after symptom onset. Of these indicators, 15 (88%) reported an increase of the number of patients with a delayed presentation to hospital services for cardiac care compared to a non-COVID-19 period. One indicator (5.9%) reported a stable number of patients with a delayed presentation, and one indicator a decrease.

### Patients’ clinical severity

Regarding the patients’ clinical severity, 136 indicators were identified from 38 articles. Most of the indicators were markers for patients with ACS, such as left ventricular ejection fraction, Killip class or biomarkers at admission. Other indicators reported on heart failure and cardiac surgery. Of these indicators, 54 (40%) reported a worse clinical condition at admission when compared with patients during the COVID-19 period, 72 indicators (53%) reported a stable clinical condition, and 10 indicators (7.4%) reported a better clinical condition.

## Diagnosis

A total of 19 indicators regarding the ‘diagnosis’ phase of the hospital cardiac care pathway were identified from 11 articles. All the indicators reported on the same category: the number of diagnostic procedures, including transthoracic and transoesophageal echocardiograms, non-invasive ischemia tests and coronary angiographies. Of these indicators, 18 (95%) reported a decrease during the COVID-19 period. One indicator (5.3%) reported a stable number of diagnostic procedures performed.

## Treatment

A total of 221 indicators regarding the ‘treatment’ phase of the cardiac care pathway were identified. These indicators were grouped into three separate categories: ‘procedure numbers’, ‘length of stay’, and ‘ACS treatment pathway times’.

### Procedure numbers

Regarding the number of treatment procedures, 79 indicators were identified from 32 articles. The indicators collected mainly concerned the number of percutaneous coronary interventions. Of these indicators, 64 (81%) reported a decrease in the number of treatment procedures performed during the COVID-19 period, and 15 indicators (19%) reported a stable number of procedures.

### Length of stay

Regarding the patients’ in-hospital length of stay, 36 indicators were identified from 25 articles. Most of the indicators reported on patients with ACS. Of these indicators, 21 (58%) reported a decrease in the length of stay during the COVID-19 period, 13 indicators (36%) reported a stable length of stay, and 2 indicators (5.6%) reported an increased length of stay.

### Acute Coronary Syndrome treatment pathway times

Regarding the treatment times for ACS care, 106 indicators were identified from 35 articles. Treatment pathway times reported in the papers were symptom-to-contact, symptom-to-door, symptom-to-diagnosis, symptom-to-balloon, contact-to-door, contact-to-balloon, door-to-ECG, door-to-balloon, ECG-to-balloon, diagnosis-to-balloon, first medical contact-to-catheter laboratory arrival, catheter-to-puncture, catheter laboratory arrival-to-balloon, puncture-to-balloon, and procedure time. Of these indicators, 65 (61%) reported an increase in ACS treatment pathway times during the COVID-19 period, 37 indicators (35%) reported stable treatment pathway times, and 4 indicators (3.8%) reported decreased treatment pathway times.

## Outpatient care

A total of 24 indicators regarding the ‘outpatient care’ phase of the cardiac care pathway were identified. These indicators were grouped into two separate categories: ‘outpatient activity’ and ‘telehealth’.

### Outpatient activity volume

Regarding outpatient activity, 18 indicators were identified from 8 articles. Most of the indicators concerned only to in-person outpatient activity. Of these indicators, 17 (94%) reported a decrease in outpatient activity volume during the COVID-19 period. One indicator (5.6%) reported a stable number.

### Telehealth

Regarding the use of telehealth, 6 indicators were identified from 6 articles. All the indicators reported an increased use of telehealth.

## Outcomes

A total of 148 indicators regarding the ‘outcomes’ phase of the cardiac care pathway were identified. These indicators were grouped into two categories: ‘outcomes and complications’ and ‘mortality rates’.

### Outcomes and complications

Regarding patients’ outcomes and complications, 88 indicators were identified from 31 articles. Most indicators were on patients with ACS, in particular ST-elevated myocardial infarctions (STEMI), such as left ventricular ejection fraction at discharge, Thrombolysis in Myocardial Infarction (TIMI) risk score after percutaneous coronary intervention and major adverse cardiovascular events. Of these indicators, 35 (40%) reported worse outcomes during the COVID-19 period, 46 indicators (52%) reported a stable outcome, and 7 (8.0%) a better outcome.

### Mortality rates

Regarding mortality rates, 60 indicators were identified from 37 articles. Forty-seven indicators reported on in-hospital mortality rate, from which 22 indicators (47%) signalled an increase, the same number (22,47%) reported a stable trend and 3 indicators (6%) signalled a decreasing trend. Five indicators reported on the 30-day death rate, from which 4 (80%) signalled an increasing trend and 1 (20%) a stable trend. There were 8 indicators reporting changes in mortality rates without any specific interval provided or with a different one.

Of all the indicators regarding mortality, 31 (52%) reported an increase in mortality rates during the COVID-19 period, 26 (43%) reported stable mortality rates, and 3 indicators (5%) show decreased mortality rates.

An overview of the trends reported by the indicators in each phase of the hospital cardiac care pathway (defined as desirable, undesirable, or stable, following clinical reasoning) is shown in Fig. 4. The trends reported by each indicator of the care pathway, by country, are available in Supplementary material S6. The studies included are available in Supplementary material S7.

**Figure 4.**
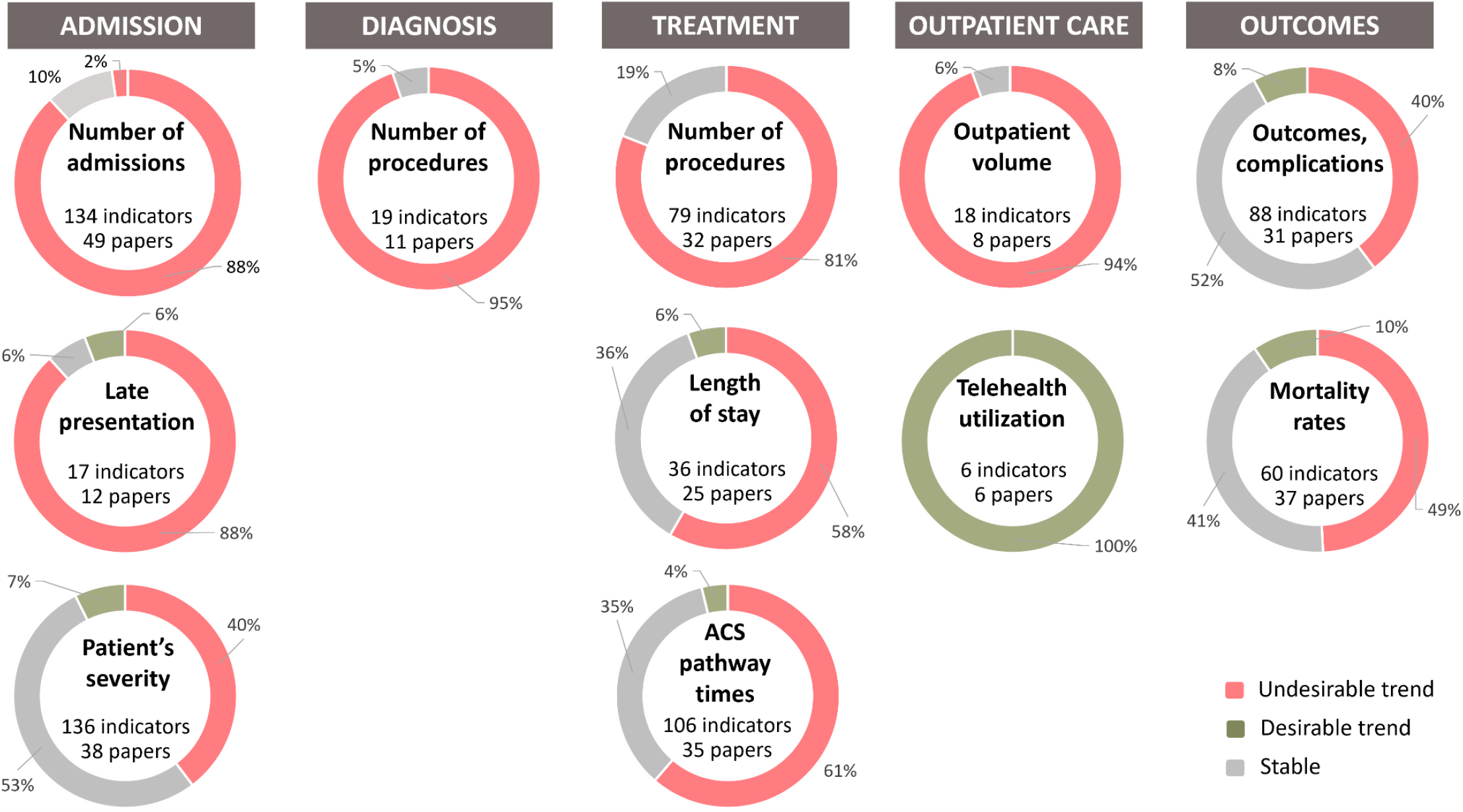
Hospital Cardiac Care Pathway Indicators’ Trends during the COVID-19 Pandemic’s early stages (Jan-Jun 2020)

## Discussion

This study aimed to provide an overview of the impact of the COVID-19 pandemic on hospital services for patients with cardiac diseases. We analysed more than 1600 indicators that were used in 94 papers, reporting on 109 different countries. Our findings show that all phases of the hospital care pathway for patients with cardiac diseases (admission, diagnosis, treatment, outpatient care and outcomes) have been, in different degrees, affected during the pandemic. Admission numbers dropped substantially, and patients arrived later and in a worse clinical condition at the hospital. The number of diagnostic and treatment procedures decreased, ACS treatment pathway times increased, and patients were discharged from the hospital after a shorter length of stay. Outpatient activity decreased, whereas the use of telehealth services increased. Finally, worse clinical outcomes and an increase in mortality rates were reported. These findings can be useful to inform clinicians and policy makers on the main areas affected in the cardiac care pathway, contributing to the monitoring and improvement of health care delivery during the current pandemic and in future unexpected crises. Additionally, these results can be helpful in planning the recovery of care for patients with cardiac diseases.

The drop in admissions to hospital services reported in almost all included articles is probably signalling healthcare avoidance, caused by patients afraid of being infected with SARS-CoV-2 in the hospital (25). Another explanation could be an actual reduction in the incidence of cardiac diseases during the pandemic. Several explanations have been opted for this reduction in incidence, such as changes in physical activity during lockdowns or a reduction of air pollutants (26-29). Patients with cardiac diseases presenting later to the hospital and arriving in a worse clinical condition than before the pandemic could also have been caused by the fear and avoidance mentioned before, with patients waiting for longer periods before seeking care.

Following the drop in admission numbers and decreased outpatient activity, a decrease in the number of diagnostic and treatment procedures were to be expected. On top of patients potentially avoiding care and a possible lower incidence rate of cardiac diseases, elective procedures and surgeries were cancelled (30). These three factors together have likely contributed to reduce procedure volume. The decreased length of hospital stay that we found in most indicators is explained by several authors by a shortage of hospital bed capacity or the physicians’ intention to minimise the risk of patients’ exposure to the virus. For instance, the European Society of Cardiology recommended that patients “should stay in the hospital for the shortest time possible” (31), and healthcare providers in the United Kingdom were advised to reduce NSTEMI inpatient stays to 36-48 hours if normal pathways could not be followed (32). The increased ACS treatment pathway times and the decreased length of stay, together, indicate that the hospital cardiac care pathway was under pressure during the COVID-19 pandemic and the delivery of care got squeezed.

The reduction in outpatient care volume we show in this study mostly comes from indicators reporting on in-patient visits. Therefore, it is probably an underestimation of the actual outpatient care provided. The change towards using telehealth shows that innovative measures were taken during the pandemic to avoid the risk of SARS-CoV-2 infection, and health systems adapted quickly to reach out to patients with other means than in-person consultations. In theory, strengthening the use of telehealth to provide continuity of care is beneficial. However, little is known about the quality of care provided and potential inequalities that may have risen from the use of telehealth (e.g., access for all patient groups).

Our findings suggest patients with cardiac conditions are showing worse outcomes and increased mortality rates during the COVID-19 pandemic. The indicators that reported changes on mortality mostly concerned short-term mortality rates (up to 30 days). An even larger impact on mortality might be found when long-term mortality rates will be assessed in later studies. These studies will have to contribute to capturing the real impact that this paper has outlined. The repercussions of care avoidance and cancelled or delayed diagnostic and treatment procedures could reveal itself in the years to come.

To our knowledge, this is the first scoping review that gives an overview of the impact of the COVID-19 pandemic on the hospital cardiac care pathway in OECD countries. The scoping review methodology gives the benefit of mapping the published literature that has come available during the early stages of the COVID-19 pandemic (January - June 2020). While earlier works assessed the impact of the pandemic on specific cardiac diseases care (33), in a specific country (34), or nation-wide, and previous authors have mentioned some of the impact revealed in this work based on earlier articles (35), this study has the strength to present an overview of performance indicators’ trends collated in a systematic way, which capture the impact throughout the hospital cardiac care pathway, and with an international scope. This methodology, while it constitutes an innovative approach, allowed to map performance indicators that could be used by countries to assess the impact of the pandemic as it evolves, in a more uniform and comparable way.

However, a scoping review has its limitations. Ideally a systematic review with meta-analyses would be performed. For the articles included in this paper this would be extremely difficult, given their different methodologies, indicators, indicators’ definitions, and comparison periods. In fact, the heterogeneity in study characteristics and indicators make it hard to compare data in a reliable way. In July 2020, the International Training Network for Healthcare Performance Intelligence Professionals (HealthPros) (36) suggested that to compare, manage and improve health systems responsiveness to the pandemic, commonly agreed-upon standardized data and indicators are necessary (37). We recommend devising a uniformly accepted set of indicators with clear definitions to use in future pandemics.

Another limitation is that only articles reporting on OECD countries were included in this scoping review. The global repercussions of the COVID-19 pandemic on hospital services for cardiac care will therefore likely be larger than reported in this paper, given the smaller capacity to handle changes in hospital care in low-income countries. On the other hand, publication bias may have played a role in portraying a more severe impact of the pandemic. Physicians might publish sooner when effects of the pandemic are being clearly noticed in their hospital. It could also be the case that the overload with clinical tasks did not allow physicians to find the time to do scientific work, which may counterbalance this limitation. Regardless, we consider our findings to be useful and signalling major trends.

Based on the included articles we are not able to provide an analysis of what caused these changes in the hospital cardiac care pathway. Being able to contextualise these results with future research will be of use for implementing measures to improve care during the current and future pandemics or disasters. Studies systematically assessing the following phases of the pandemic are necessary to evaluate whether the cardiac patients’ management improved. We also welcome studies on the quality and access of telemedicine. It would be relevant to study if and how this could improve quality and access of services delivery for the better, also in non-crisis times.

In conclusion, these results signal the hospital services delivery process for cardiac care came under pressure in the first half of 2020, and all phases of the hospital cardiac care pathway were affected. Lessons should be learnt, and steps taken to be able to safeguard the continuity of care during the ongoing COVID-19 pandemic, and in future crises. Furthermore, to guide the decisions of health system actors on the implementation of measures to ensure the continuation of essential care during future crises, fostering the use of an international standardised set of indicators is paramount, making optimal use of existing health information infrastructures.

## Data Availability

All data produced are available online in Zenodo.org, at https://dx.doi.org/10.5281/zenodo.5745755.

## Supplementary data

Supplementary material S1 – Preferred Reporting Items for Systematic reviews and Meta-Analyses extension for Scoping Reviews (PRISMA-ScR) Checklist

Supplementary material S2 – Search strategy Supplementary material S3 – Data Extraction Form

Supplementary material S4 – Collected and grouped indicators

Supplementary material S5 – Characteristics of the studies included in the review, and from which indicators were extracted and collated

Supplementary material S6 – Number of indicators and indicators’ trends reported by each indicator, presented by country

Supplementary material S7 – Additional references: studies included in the review, and from which indicators were extracted and collated

## Acknowledgements

MdL and ASC contributed equally as co-first authors to the development and execution of this study. The authors wish to thank Wichor Bramer from the Erasmus MC Medical Library for developing and updating the search strategies used in this study.

## Ethics approval and consent to participate

Not required.

## Data availability

The data underlying this article are available in Zenodo.org, at https://dx.doi.org/10.5281/zenodo.5745755 (38).

## Authors’ contributions

All authors contributed to conceptualize the study. ASC, MdL and OBF performed the data collection. MdL performed the data analysis with collaboration and revision by ASC. MdL and ASC drafted the article. All authors provided feedback and contributed to revising the manuscript. All authors approved the final version of the manuscript.

## Key points

- This study aims to assess the impact of the pandemic on hospital care for cardiac patients in OECD countries from January to June-2020, by collecting and categorizing performance indicators that reported changes in health care use and quality.
- This study used a scoping review methodology, we analysed more than 1600 indicators from 94 papers, along with the trends reported by those indicators, regarding the different phases of the hospital cardiac care pathway: admission, diagnosis, treatment, outpatient care, outcomes.
- All phases of the hospital cardiac care pathway have been, in different degrees, affected, since admission numbers dropped substantially, patients arrived later and in a worse clinical condition at the hospital, while the number of diagnostic and treatment procedures decreased.
- Our findings also show that acute coronary syndromes’ treatment pathway times increased, patients were discharged from the hospital after a shorter length of stay, outpatient activity decreased, the use of telehealth services increased, and worse clinical outcomes were reported.
- While these findings can be useful to inform on the areas affected in the early phase of the current pandemic, fostering the use of an international standardised set of indicators is paramount to guide the decisions of health system actors to ensure the continuation of essential care during this pandemic and in future crises, making optimal use of existing health information infrastructures.

