## Supplementary data for "The impact of the COVID-19 pandemic on hospital services for patients with cardiac diseases: a scoping review"

**Supplementary material S1 - Preferred Reporting Items for Systematic reviews and Meta-Analyses extension for Scoping Reviews (PRISMA-ScR) Checklist**

| SECTION | ITEM | PRISMA-ScR CHECKLIST ITEM | REPORTED ON PAGE # |
|---|---|---|---|
| **TITLE** | | | |
| Title | 1 | Identify the report as a scoping review. | Title Page |
| **ABSTRACT** | | | |
| Structured summary | 2 | Provide a structured summary that includes (as applicable): background, objectives, eligibility criteria, sources of evidence, charting methods, results, and conclusions that relate to the review questions and objectives. | 2 |
| **INTRODUCTION** | | | |
| Rationale | 3 | Describe the rationale for the review in the context of what is already known. Explain why the review questions/objectives lend themselves to a scoping review approach. | 3 |
| Objectives | 4 | Provide an explicit statement of the questions and objectives being addressed with reference to their key elements (e.g., population or participants, concepts, and context) or other relevant key elements used to conceptualize the review questions and/or objectives. | 3 |
| **METHODS** | | | |
| Protocol and registration | 5 | Indicate whether a review protocol exists; state if and where it can be accessed (e.g., a Web address); and if available, provide registration information, including the registration number. | |
| Eligibility criteria | 6 | Specify characteristics of the sources of evidence used as eligibility criteria (e.g., years considered, language, and publication status), and provide a rationale. | 5 |
| Information sources* | 7 | Describe all information sources in the search (e.g., databases with dates of coverage and contact with authors to identify additional sources), as well as the date the most recent search was executed. | 4 |
| Search | 8 | Present the full electronic search strategy for at least 1 database, including any limits used, such that it could be repeated. | 4 |
| Selection of sources of evidence† | 9 | State the process for selecting sources of evidence (i.e., screening and eligibility) included in the scoping review. | 6 |
| Data charting process‡ | 10 | Describe the methods of charting data from the included sources of evidence (e.g., calibrated forms or forms that have been tested by the team before their use, and whether data charting was done independently or in duplicate) and any processes for obtaining and confirming data from investigators. | 6 |
| Data items | 11 | List and define all variables for which data were sought and any assumptions and simplifications made. | 6 |
| Critical appraisal of individual sources of evidence§ | 12 | If done, provide a rationale for conducting a critical appraisal of included sources of evidence; describe the methods used and how this information was used in any data synthesis (if appropriate). | Not applicable |
| Synthesis of results | 13 | Describe the methods of handling and summarizing the data that were charted. | 7 |

| SECTION | ITEM | PRISMA-ScR CHECKLIST ITEM | REPORTED ON PAGE # |
|---|---|---|---|
| **RESULTS** | | | |
| Selection of sources of evidence | 14 | Give numbers of sources of evidence screened, assessed for eligibility, and included in the review, with reasons for exclusions at each stage, ideally using a flow diagram. | 8 |
| Characteristics of sources of evidence | 15 | For each source of evidence, present characteristics for which data were charted and provide the citations. | 8 |
| Critical appraisal within sources of evidence | 16 | If done, present data on critical appraisal of included sources of evidence (see item 12). | Not applicable |
| Results of individual sources of evidence | 17 | For each included source of evidence, present the relevant data that were charted that relate to the review questions and objectives. | 8-12 |
| Synthesis of results | 18 | Summarize and/or present the charting results as they relate to the review questions and objectives. | 8-12 |
| **DISCUSSION** | | | |
| Summary of evidence | 19 | Summarize the main results (including an overview of concepts, themes, and types of evidence available), link to the review questions and objectives, and consider the relevance to key groups. | 13 |
| Limitations | 20 | Discuss the limitations of the scoping review process. | 15 |
| Conclusions | 21 | Provide a general interpretation of the results with respect to the review questions and objectives, as well as potential implications and/or next steps. | 14-15 |
| **FUNDING** | | | |
| Funding | 22 | Describe sources of funding for the included sources of evidence, as well as sources of funding for the scoping review. Describe the role of the funders of the scoping review. | 17 |

JBI = Joanna Briggs Institute; PRISMA-ScR = Preferred Reporting Items for Systematic reviews and Meta-Analyses extension for Scoping Reviews.

* Where *sources of evidence* (see second footnote) are compiled from, such as bibliographic databases, social media platforms, and Web sites.

† A more inclusive/heterogeneous term used to account for the different types of evidence or data sources (e.g., quantitative and/or qualitative research, expert opinion, and policy documents) that may be eligible in a scoping review as opposed to only studies. This is not to be confused with *information sources* (see first footnote).

‡ The frameworks by Arksey and O'Malley (6) and Levac and colleagues (7) and the JBI guidance (4, 5) refer to the process of data extraction in a scoping review as data charting.

§ The process of systematically examining research evidence to assess its validity, results, and relevance before using it to inform a decision. This term is used for items 12 and 19 instead of "risk of bias" (which is more applicable to systematic reviews of interventions) to include and acknowledge the various sources of evidence that may be used in a scoping review (e.g., quantitative and/or qualitative research, expert opinion, and policy document).

*From:* Tricco AC, Lillie E, Zarin W, O'Brien KK, Colquhoun H, Levac D, et al. PRISMA Extension for Scoping Reviews (PRISMAScR): Checklist and Explanation. Ann Intern Med. 2018;169:467–473. ***doi: 10.7326/M18-0850***.

#### Supplementary material 2 – Search strategy

| Database searched | via | Years of coverage | Records | Records after duplicates removed |
|---|---|---|---|---|
| Embase | Embase.com | 1971 - Present | 4282 | 4206 |
| Medline ALL | Ovid | 1946 - Present | 4429 | 2071 |
| **Total** | | | **8711** | **6277** |

**Embase.com 4282**

(pandemic/mj/exp OR 'natural disaster'/mj/exp OR disaster/mj/exp OR hurricane/mj/exp OR earthquake/mj/de OR 'coronavirus disease 2019'/mj/de OR 'Severe acute respiratory syndrome coronavirus 2'/mj/de OR (pandemic* OR disaster* OR hurricane* OR flood OR flooding OR earthquake* OR coronavirus-disease-2019 OR covid19 OR covid-19 OR sars-cov-2 OR 2019-novel-coronavirus OR 2019-ncov OR lockdown OR lock-down OR typhoon* OR (natural NEAR/3 hazard*)):ti) AND ('non communicable disease'/de OR neoplasm/exp OR 'malignant neoplasm'/de OR 'cancer patient'/exp OR 'heart disease'/exp OR cardiology/de OR 'neurologic disease'/de OR 'cancer screening'/de OR 'cancer surgery'/de OR (('primary medical care'/exp OR 'general practitioner'/de OR 'general practice'/de OR 'family medicine'/de OR 'emergency ward'/exp OR 'emergency care'/de) AND ('health care utilization'/exp OR consultation/de)) OR 'diabetes mellitus'/exp OR 'rheumatic disease'/exp OR 'mental health service'/de OR 'cardiovascular disease'/exp OR 'cerebrovascular disease'/exp OR 'chronic kidney failure'/exp OR 'obstructive airway disease'/exp OR 'chronic respiratory tract disease'/de OR 'cerebrovascular accident'/exp OR hypertension/exp OR 'chronic disease'/de OR 'neurologic disease'/exp OR (((non-communicab* OR noncommunicab*) NEAR/3 disease) OR cancer OR ((heart OR cardiovascul* OR cerebrovascul*) NEAR/3 (disease* OR patient* OR emergenc*)) OR cardiolog* OR neurolog* OR oncolog* OR ((((emergency OR acute) NEAR/3 (ward* OR care OR department*)) OR (general NEXT/1 (practi*)) OR ((primary OR family) NEXT/2 (healthcare* OR care OR doctor*))) AND (utilization* OR utilisation* OR delay* OR time OR visit* OR consultation* OR barrier* OR access* OR challenge* OR vulnerab* OR attendan* OR hesita*)) OR diabet* OR rheumat* OR (mental NEAR/3 (health-care* OR health-service* OR healthcare*)) OR (chronic NEAR/3 (kidney OR renal OR respirator* OR lung* OR pulmonar*) NEAR/3 (failure OR disease*)) OR asthma* OR bronchitis OR ((lung* OR pulmonar*) NEAR/3 (emphysem*)) OR ((cerebrovascul* OR cerebro-vascul*) NEAR/3 accident*) OR stroke OR hypertens* OR chronic-disease* OR (surg* NEAR/3 emergenc*)):ab,ti) AND ('disease exacerbation'/mj/de OR 'recurrent disease'/mj/de OR 'avoidance behavior'/mj/de OR 'recurrence risk'/mj/de OR 'undiagnosed disease'/mj/de OR 'diagnostic error'/mj/de OR 'diagnostic delay'/mj/de OR 'delayed diagnosis'/mj/de OR 'therapy delay'/mj/de OR 'health care utilization'/mj/de OR mortality/mj/exp OR morbidity/mj/exp OR 'health care quality'/mj/exp OR screening/mj OR 'mass screening'/exp/mj OR 'screening test'/mj OR 'palliative therapy'/exp/mj OR 'health care delivery'/mj OR 'health care access'/exp/mj OR 'surgical volume'/mj OR rehabilitation/exp/mj OR 'performance indicator'/de OR (impact* OR influence* OR affect* OR effect* OR exacerbate* OR (Disease NEAR/3 progression*) OR recur* OR avoid* OR postpon* OR post-pon* OR implication* OR ((diagnos* OR therap* OR treatment* OR hospitali* OR services OR visit*) NEAR/3 (error* OR delay* OR fewer* OR drop OR decline* OR continuit* OR decrease* OR increase* OR reduc*)) OR ((health-care OR healthcare) NEAR/3 (utilizat* OR utilisat* OR use)) OR mortalit* OR morbidit* OR ((health-care OR healthcare) NEAR/3 (quality)) OR screening OR (service* NEAR/3 ( disrupt* OR continu*)) OR palliat* OR unmet OR rehabilitation*):ti OR (((performance* OR outcome*) NEAR/3 (indicator*)) OR ((health-care OR healthcare) NEAR/3 (access* OR deliver* OR output*)) OR ((surgical OR surgeries OR procedures) NEAR/3 (volum* OR number*))):ab,ti) NOT (model/exp/mj OR (model*):ti) NOT ([conference abstract]/lim AND [2000-2019]/py) NOT ([animals]/lim NOT [humans]/lim) NOT ('health care personnel'/exp/mj OR pregnancy/exp/mj OR 'pregnant woman'/mj OR (((COVID-19 OR COVID19 OR coronavirus* OR corona-virus* OR SARS-CoV-2) NEAR/3 (outcome* OR case* OR patient* OR pneumoni* OR progression* OR prognos* OR mortalit* OR fatal* OR sever* OR vaccine*)) OR pregnan* OR ((healthcare OR health-care OR medical) NEAR/3 (personnel* OR staff OR worker* OR workforce* OR work-force*)) OR doctor* OR nurse* OR physician*):ti OR ((COVID-19 OR COVID19 OR coronavirus* OR corona-virus* OR SARS-CoV-2) NEAR/3 (mortalit* OR outcome* OR fatal*)):ab) NOT ((child/exp NOT adult/exp) OR (pediatr* OR paediatr* OR child* OR infan* OR adolescen*):ti) NOT ('practice guideline'/de OR (guideline*):ti)

**Medline ALL Ovid 4429**

(*Pandemics/ OR *Natural Disasters/ OR *Cyclonic Storms/ OR *Earthquakes/ OR *COVID-19/ OR *SARS-CoV-2/ OR (pandemic* OR disaster* OR hurricane* OR flood OR flooding OR earthquake* OR coronavirus-disease-2019 OR covid19 OR covid-19 OR sars-cov-2 OR 2019-novel-coronavirus OR 2019-ncov OR lockdown OR lock-down OR typhoon* OR (natural ADJ3 hazard*)).ti.) AND (Noncommunicable Diseases/ OR exp Neoplasms/ OR exp Heart Diseases/ OR Cardiology/ OR exp Nervous System Diseases/ OR Early Detection of Cancer/ OR exp Diabetes Mellitus/ OR exp Rheumatic Diseases/ OR exp Mental Health Services/ OR exp Cardiovascular Diseases/ OR exp Cerebrovascular Disorders/ OR exp Kidney Failure, Chronic/ OR Lung Diseases, Obstructive/ OR exp Stroke/ OR exp Hypertension/ OR Chronic Disease/ OR ((exp Emergency Medical Services/ OR exp Emergency Treatment/ OR General Practitioners/ OR General Practice/ OR Family Practice/) AND (Patient Acceptance of Health Care/ OR "Referral and Consultation"/)) OR (((non-communicab* OR noncommunicab*) ADJ3 disease) OR cancer OR ((heart OR cardiovascul* OR cerebrovascul*) ADJ3 (disease* OR patient* OR emergenc*)) OR cardiolog* OR neurolog* OR oncolog* OR ((((emergency OR acute) ADJ3 (ward* OR care OR department*)) OR (general ADJ (practi*)) OR ((primary OR family) ADJ2 (healthcare* OR care OR doctor*))) AND (utilization* OR utilisation* OR delay* OR time OR visit* OR consultation* OR barrier* OR access* OR challenge* OR vulnerab* OR attendan* OR hesita*)) OR diabet* OR rheumat* OR (mental ADJ3 (health-care* OR health-service* OR healthcare*)) OR (chronic ADJ3 (kidney OR renal OR respirator* OR lung* OR pulmonar*) ADJ3 (failure OR disease*)) OR asthma* OR bronchitis OR ((lung* OR pulmonar*) ADJ3 (emphysem*)) OR ((cerebrovascul* OR cerebro-vascul*) ADJ3 accident*) OR stroke OR hypertens* OR chronic-disease* OR (surg* ADJ3 emergenc*)).ab,ti.) AND (*Disease Progression/ OR exp * Recurrence/ OR *Undiagnosed Diseases/ OR exp *Diagnostic Errors/ OR *Delayed Diagnosis/ OR * Time-to-Treatment/ OR *Patient Acceptance of Health Care/ OR exp *Mortality/ OR exp *Morbidity/ OR exp *Quality of Health Care/ OR *Early Detection of Cancer/ OR *Mass Screening/ OR *Palliative Care/ OR exp *Delivery of Health Care/ OR *Health Services Accessibility/ OR *Rehabilitation/ OR (impact* OR influence* OR affect* OR effect* OR exacerbate* OR (Disease ADJ3 progression*) OR recur* OR avoid* OR postpon* OR post-pon* OR implication* OR ((diagnos* OR therap* OR treatment* OR hospitali* OR services OR visit*) ADJ3 (error* OR delay* OR fewer* OR drop OR decline* OR continuit* OR decrease* OR increase* OR reduc*)) OR ((health-care OR healthcare) ADJ3 (utilizat* OR utilisat* OR "use")) OR mortalit* OR morbidit* OR ((health-care OR healthcare) ADJ3 (quality)) OR screening OR (service* ADJ3 ( disrupt* OR continu*)) OR palliat* OR unmet OR rehabilitation*).ti. OR (((performance* OR outcome*) ADJ3 (indicator*)) OR ((health-care OR healthcare) ADJ3 (access* OR deliver* OR output*)) OR ((surgical OR surgeries OR procedures) ADJ3 (volum* OR number*))).ab,ti.) NOT (exp * Models, Theoretical/ OR model*:ti) NOT (exp animals/ NOT humans/) NOT (exp * Health Personnel/ OR exp * Attitude of Health Personnel / OR exp * pregnancy/ OR * pregnant woman/ OR (((COVID-19 OR COVID19 OR coronavirus* OR corona-virus* OR SARS-CoV-2) ADJ3 (outcome* OR case* OR patient* OR pneumoni* OR progression* OR prognos* OR mortalit* OR fatal* OR sever* OR vaccine*)) OR pregnan* OR ((healthcare OR health-care OR medical) ADJ3 (personnel* OR staff OR worker* OR workforce* OR work-force*)) OR doctor* OR nurse* OR physician*).ti. OR ((COVID-19 OR COVID19 OR coronavirus* OR corona-virus* OR SARS-CoV-2) ADJ3 (mortalit* OR outcome* OR fatal*)).ab.) NOT (((exp child/ OR exp infant/) NOT exp adult/) OR (pediatr* OR paediatr* OR child* OR infan* OR adolescen*).ti.) NOT (Practice Guideline/ OR Practice Guidelines as Topic/ OR (guideline*).ti.)

#### Supplementary material 3 – Data Extraction Form

##### Form 1: General study characteristics

| | |
|---|---|
| Article ID | Number identifying each paper |
| Article source | Systematic search |
| | Reference mining |
| Clinical area | |
| Disease/care delivered | |
| Country | |
| Study design | Randomized controlled trial |
| | Prospective cohort |
| | Retrospective cohort |
| | Case-control |
| | Cross-sectional |
| | Case report |
| | Systematic review |
| | Meta-analysis |
| | Scoping review |
| | Survey |
| Peer-reviewed | Yes / No |
| Excluded: (with reason for exclusion) | Yes (reason) / No |
| Time period studied | |
| Comparison period | 2020 vs. 2019 |
| | 2020: pre- versus post-COVID |
| | 2020 vs. previous years |
| Article title | |
| Year of publication | |
| Journal | |
| Authors | |
| Paper URL | |

| |
|---|
| Language |
| Abstract |

**Form 2: Indicator related information**

| | |
|---|---|
| Paper ID | Number identifying each paper |
| Clinical area | |
| Disease/care delivered | |
| Country | |
| Data source | Population-level data |
| | Adverse incident data |
| | Clinical data |
| | Costing data |
| | Claims data |
| | Survey data |
| | Registry data |
| | Prescribing data |
| | Claims data |
| Database | |
| Sample size | Number of centers |
| | Number of patients/procedures/responses/events |
| Indicator title | |
| Self-reported | Yes |
| | No |
| Numerator (inclusion criteria) | |
| Numerator (exclusion criteria) | |
| Denominator | |
| Indicator results from statistical model | Yes |
| | No |
| Periods being compared | 2020 vs. 2019 |

| | |
|---|---|
| | 2020: pre- versus post-COVID |
| | 2020 vs. previous years |
| Trend reported by indicator | Increase |
| | Stable |
| | Decrease |
| Numerical value before pandemic | |
| Numerical value during pandemic | |
| Units relating to numerical value | (eg: ng/L, days, admissions) |
| Magnitude (%) | *Computed as percent change:*<br>((value during pandemic - value before pandemic) /<br>(value before pandemic)) * 100 |
| Main results (if reported) | Relative risk ratio |
| | Odds ratio |
| | Incidence rate ratio |
| Other results reported (free field) | |
| Delivery of care pathway | Access / admission |
| | Diagnostic |
| | Treatment |
| | Follow-up / Outpatient care |
| | Other |
| | Outcomes |
| Dimensions of performance | Access |
| | Quality – effectiveness |
| | Quality – safety |
| | Quality – patient-centredness |
| Donabedian's framework: | Structure |
| | Process |
| | Outcome |

#### Supplementary material 4 – Collected and grouped indicators

##### Admission

| Admission numbers | |
|---|---|
| **Indicator name** | **Number of papers** |
| Cardiovascular hospitalizations | 3 |
| Cardiology emergency admissions | 2 |
| Elective cardiology admissions | 1 |
| Admissions for acute cardiac events | 1 |
| Admitted to emergency department with cardiovascular symptoms | 1 |
| Admissions to cardiology ward / coronary care unit | 1 |
| Acute coronary syndrome (ACS) hospital admissions | 13 |
| Acute myorcardial infarction (STEMI/NSTEMI) hospital admissions | 8 |
| NSTEMI admissions | 17 |
| STEMI admissions | 24 |
| Unstable angina admissions | 8 |
| Heart failure hospital admissions | 13 |
| Admissions for cardiac arrhythmia | 6 |
| Admissions for out of hospital cardiac arrest | 1 |
| Valvular heart disease admissions | 1 |
| Myocarditis, pericarditis admissions | 1 |
| Number of hospitalizations for device malfunctioning | 1 |
| Threshold for hospitalization of heart failure patients | 1 |

| Delayed presentation | |
|---|---|
| **Indicator name** | **Number of papers** |
| ACS - Late presenting | 1 |
| STEMI - late presenting | 11 |
| NSTEMI - late presenting | 2 |
| AMI - self-presenting to hospital | 4 |

| Patient's clinical condition | |
|---|---|
| **Indicator name** | **Number of papers** |
| ACS - Left ventricular ejection fraction | 2 |
| ACS - Admission troponin | 2 |
| ACS - Admission CK | 1 |
| ACS - GRACE-score | 1 |
| ACS - Number of diseased vessels | 3 |
| ACS - Charlson comorbidity index | 1 |
| ACS - Cardiogenic shock | 2 |
| ACS - Life threatening arrhytmia | 2 |
| ACS - Intubation | 1 |
| ACS - Inotropic support | 1 |
| ACS - Resuscitation | 1 |
| ACS - Hemodynamic support | 1 |
| ACS - Invasive ventilation | 1 |
| AMI - Cardiogenic shock | 1 |
| STEMI patients admitted to ICU/CCU | 2 |
| STEMI - Ejection fraction at presentation | 9 |
| STEMI - Killip class at presentation | 7 |
| STEMI - Cardiogenic shock | 8 |
| STEMI - Cardiac arrest | 8 |
| STEMI - Asystole | 1 |
| STEMI - Ventricular fibrillation | 2 |
| STEMI - Mechanical ventilation | 2 |
| STEMI - Troponin | 5 |
| STEMI - LDH | 2 |
| STEMI - CK | 2 |
| STEMI - NTproBNP | 1 |
| STEMI - GRACE-score | 1 |
| STEMI - baseline TIMI-flow | 7 |
| STEMI - thrombosis degree | 1 |
| STEMI - Collateral circulation | 1 |
| STEMI - Number of vessels | 8 |
| STEMI - Presence of new Q-waves on the earliest ECG | 1 |
| STEMI - Atrial fibrillation | 2 |
| NSTEMI patients admitted to CCU | 1 |
| NSTEMI - GRACE-score | 1 |
| NSTEMI - Troponin | 1 |
| Heart failure - NYHA class | 2 |
| Heart failure - Charlson comorbidity index | 3 |
| Heart failure - LVEF | 2 |
| Heart failure - NT-proBNP | 2 |
| Heart failure - admission to ICU | 3 |
| Heart failure - SBP < 100mmHg on admission | 1 |
| Heart failure - eGFR < 30 | 1 |
| Acute heart failure patients admitted to ICU | 1 |
| Cardiac surgery - LVEF | 1 |
| Cardiac surgery - Preoperative EuroSCORE II | 1 |
| Cardiac surgery - patients admitted with a critical preoperative status | 1 |
| Patients admitted in cardiac intensive care unit - LVEF | 1 |

#### Diagnostics

| Diagnostics | |
|---|---|
| **Indicator name** | **Number of papers** |
| Number of angiographies | 6 |
| Number of SPECT-MPI | 1 |
| Number of cardiac imaging procedures | 3 |
| Number of non-invasive ischemia tests | 2 |
| Number of elective diagnostic catheterisation | 1 |
| Number of acute cardiac tests | 1 |
| Number of ECG's | 1 |
| Number of NT-proBNP blood tests in primary and secondary care | 1 |
| Number of dobutamine stress echocardiagram | 1 |

### Treatment

| Procedure numbers | |
|---|---|
| **Indicator name** | **Number of papers** |
| Number of cardiac procedures performed | 4 |
| Number of PCI procedures | 22 |
| Number of transcatheter aortic valve replacements | 4 |
| Number of structural heart disease procedures | 3 |
| Number of mitraclip | 1 |
| Number of CABG procedures | 5 |
| Number of pacemaker/defibrillator implantations | 4 |
| Number of electrophysiology procedures | 2 |
| Number of cardioversions | 2 |
| Number of ablations | 1 |
| Number of cardiovascular surgeries | 4 |
| Heart failure - % of pts who underwent cardiac resynchronization therapy device implantation | 1 |
| Number of invasive coronary procedures because of ACS | 1 |
| Transplant cardiac activity | 1 |

| Length of stay | |
|---|---|
| **Indicator name** | **Number of papers** |
| Patients admitted to cardiology departments - length of stay | 1 |
| Patients admitted to cardiac intensive care unit/ICU - length of stay | 5 |
| Acute cardiovascular conditions - length of stay | 1 |
| Heart failure patients - average length of stay | 3 |
| Acute coronary syndrome (ACS) - length of stay | 4 |
| Acute myocardial infarction - length of stay | 5 |
| STEMI - length of stay | 11 |
| NSTEMI - length of stay | 3 |
| Postprocedure same-day discharge | 1 |

| ACS pathway times | |
|---|---|
| **Indicator name** | **Number of papers** |
| Symptom-to-contact | 17 |
| Symptom-to-door | 9 |
| Symptom-to-diagnosis | 2 |
| Symptom-to-balloon (ischaemia time) | 14 |
| Contact-to-door | 3 |
| Contact-to-balloon | 10 |
| Door-to-ECG | 1 |
| Door-to-balloon | 24 |
| ECG-to-balloon | 2 |
| Diagnosis-to-balloon | 2 |
| First medical contact to cath lab arrival | 1 |
| Cath-to-puncture | 1 |
| Cath to balloon | 2 |
| Puncture-to-balloon | 1 |
| Procedure time | 3 |

### Outpatient care

| Outpatient activity & telehealth | |
|---|---|
| **Indicator name** | **Number of papers** |
| Outpatient activity | 4 |
| Referral to cardiac rehabilitation | 3 |
| Number of referrals to cardiology outpatient clinics | 1 |
| Number of in-person outpatient visits | 2 |
| Cancelled outpatient cardiology visits | 2 |
| Patients reporting that HF appointments were negatively impacted during the lockdown period | 1 |
| Outpatient care converted to tele-health | 2 |
| Proportion of telehealth visits | 4 |

#### Outcomes

| Patient outcomes and complications | |
|---|---|
| **Indicator name** | **Number of papers** |
| LVEF <40% at discharge / <35% at discharge | 3 |
| Post revascularization left ventricular ejection fraction | 2 |
| Post infarction left ventricular ejection fraction | 2 |
| Left ventricular ejection fraction at discharge | 4 |
| Occurence of stent thrombosis | 5 |
| Cardiogenic shock / mechanical complication in STEMI | 1 |
| In-hospital re-PCI | 2 |
| Major adverse cardiovascular event (MACE) | 5 |
| Rates of emergency department / hospitalization 30 days post in-person visits | 1 |
| Cardiogenic shock / CPR after cardiovasular emergencies | 1 |
| ACS/STEMI patients developing mechanical complication | 4 |
| Troponin peak value | 6 |
| TIMI flow after PCI | 11 |
| STEMI patients - Pericardial effusion | 1 |
| STEMI patients - Bleeding requiring blood transfusion | 2 |
| STEMI patients - AKI / Renal failure / Dialysis | 3 |
| STEMI - Left ventricular aneurysm | 1 |
| Reinfarction | 3 |
| STEMI - Malignant arrhytmia | 1 |
| STEMI - Cardiogenic shock | 1 |
| STEMI - Target vessel urgent revascularization | 1 |
| STEMI - Stroke | 2 |
| STEMI patients - LV thrombus | 2 |
| STEMI - Procedural success | 1 |
| STEMI - Coronary perforation | 1 |
| STEMI - Retroperitoneal bleed | 1 |
| STEMI - Thrombo embolism | 1 |
| STEMI - Mechanical ventilation | 1 |
| STEMI - Inotropic support | 1 |
| STEMI - Mechanical circulatory support | 1 |
| STEMI - Sepsis | 1 |
| STEMI - Post PCI ECG, ST resolution >70% | 1 |
| STEMI - Post PCI CK | 1 |
| Number of deaths due to infections after PCI | 1 |
| AMI - NTproBNP | 1 |
| STEMI - TIMI frame count <23 | 1 |
| 30-day hospital readmission rate for heart failure | 1 |
| Rates of ED/hospitalization 30 days post In-person visits | 1 |

| Mortality rates | |
|---|---|
| **Indicator name** | **Number of papers** |
| Intra-hospital cardiovascular mortality | 2 |
| In-hospital mortality of patients admitted to cardiology departments | 1 |
| Cardiac mortality | 2 |
| In-hospital mortality rate of acute coronary syndrome (STEMI, NSTEMI, unstable angina) | 5 |
| In-hospital mortality rate of acute myocardial infarction (STEMI, NSTEMI) | 4 |
| In-hospital mortality rate of NSTEMI | 2 |
| In-hospital mortality rate of STEMI | 16 |
| 7-day mortality AMI patients | 1 |
| 30-day mortality AMI patients | 1 |
| 30-day death rate after PCI | 1 |
| In-hospital mortality in patients admitted for cardiac catherization/PCI | 3 |
| Postdischarge mortality up to 30 days after PCI | 1 |
| All-cause mortality STEMI patients | 1 |
| Rate of death at the emergency department for cardiovascular diseases | 1 |
| Death caused by ischemic heart disease | 1 |
| In-hospital death in patients admitted to CCU | 1 |
| In-hospital mortality among cardiac surgery recipients | 1 |
| Short-term (30 day) case mortality of patients diagnosed with acute HF | 1 |
| Death caused by heart failure | 1 |
| In-hospital mortality rate of heart failure patients | 4 |

#### Supplementary material 5 – Characteristics of the studies included in the review, and from which indicators were extracted and collated

| **Reference Number** (document with references extra) | **COUNTRY** | **Study design** | **Time period studied - Start** | **Time period studied - End** | **Comparison 2020 vs. 2019** | **Comparison 2020: pre / post Covid** | **Comparison 2020 vs. previous years** |
|---|---|---|---|---|---|---|---|
| 1 | Canada | Prospective + retrospective cohort | 01-03-2020 | 19-04-2020 | 2019: same period | | |
| 2 | Multiple countries (a) | Retrospective cohort | 12-03-2020 | 15-04-2020 | | 01-12-2019 to 11-03-2020 | 12-2018 to 04-2019 |
| 3 | United Kingdom | Retrospective cohort | 23-03-2020 | 10-05-2020 | | 2018-2019 VS before 1st case: 28-10-2019 to 02-02-2020; 2018-2019 VS between 1stcase and lockdown: 03-02-2020 to 22-03-2020 | 2018-2019: corresponding dates |
| 4 | US | Retrospective cohort | 01-01-2020 | 31-03-2020 | 2019: same period | | |
| 5 | Switzerland | Retrospective cohort | 01-01-2020 | 28-02-2020 | 2019: same period | Mar-Apr 2020 | |
| 6 | Germany | Retrospective cohort | 13-03-2020 | 30-04-2020 | 2019: same period | 01-01-2020 to 12-03-2020 | |
| 7 | United Kingdom | Retrospective cohort | 02-03-2020 | 19-04-2020 | | | 2017-2019: same period |
| 8 | Israel | Retrospective cohort | 01-02-2020 | 30-06-2020 | | | 01-07-2018 to 31-01-2020 |
| 9 | Austria | Retrospective cohort | 16-03-2020 | 26-04-2020 | | 03-02-2020 to 15-03-2020 | 2016-2019: same period |
| 10 | Denmark | Prospective cohort | 13-03-2020 | 16-10-2020 | 2019: same period | | |

| | | | | | | | |
|---|---|---|---|---|---|---|---|
| 11 | Belgium | Retrospective cohort | 13-03-2020 | 03-04-2020 | | | 2017-2019: same period |
| 12 | Italy | Retrospective cohort | 20-02-2020 | 20-04-2020 | 2019: same period | | |
| 13 | Italy | Retrospective cohort | 20-02-2020 | 31-03-2020 | 2019: same period | 01-01-2020 to 19-02-202 | |
| 14 | United Kingdom | Retrospective cohort | 03-03-2020 | 27-04-2020 | | 07-01-2020 to 02-03-2020 | |
| 15 | Turkey | Prospective cohort | 17-04-2020 | 02-05-2020 | | | 2018: 1 to 15-11-2018 |
| 16 | Italy | Retrospective cohort | 01-03-2020 | 30-04-2020 | 2019: same period | | 2015-2019: same period |
| 17 | United Kingdom | Retrospective cohort | 21-04-2020 | 20-05-2020 | | 21-03-2020 to 20-04-2020 (also lockdown); 21-02 to 20-03-2020 (transition period); 21-01-2020 to 20-02-2020 (baseline) | |
| 18 | Italy | Retrospective cohort | 10-03-2020 | 10-04-2020 | 2019: same period | | |
| 19 | Italy | Retrospective cohort | 24-02-2020 | 29-03-2020 | 2019: same period | 8 weeks before | |
| 20 | Poland | Retrospective cohort | 09-03-2020 | 16-04-2020 | 2019: same period | | |
| 21 | Germany | Retrospective cohort | 01-03-2020 | 21-04-2020 | 2019: same period | | 2017-2019: same period |
| 22 | US | Retrospective cohort | 23-02-2020 | 28-03-2020 | | 29-03-2020 to 16-05-2020 | 30-12-2018 to 22-02-2020 |
| 23 | Italy | Prospective cohort | 21-02-2020 | 01-04-2020 | 2019: same period | | 2018: same period |
| 24 | Switzerland | Retrospective cohort | 17-02-2020 | 14-04-2020 | 2019: same period | | |
| 25 | Greece | Prospective cohort | 01-02-2020 | 30-06-2020 | | 01-01-2019 to 31-01-2020 | |
| 26 | Turkey | Retrospective cohort | 10-03-2020 | 30-04-2020 | | | 2015-2019: same period |
| 27 | Germany | Retrospective cohort | 01-03-2020 | 01-04-2020 | 2019: same period | 01-01-2020 to 12-03-2020 | |

| | | | | | | | |
|---|---|---|---|---|---|---|---|
| 28 | Germany | Retrospective cohort | 13-03-2020 | 10-09-2020 | 2019: same period | | |
| 29 | United Kingdom | Retrospective cohort | 23-03-2020 | 24-05-2020 | 2019: weekly/monthly average | 01-01-2019 to 22-03-2020 | |
| 30 | US | Survey | 30-04-2020 | 13-05-2020 | | "Compared to pre-Covid" | |
| 31 | Spain | Retrospective cohort | 12-03-2020 | 20-04-2020 | | 01-02-2020 to 11-03-2020 | |
| 32 | Germany | Retrospective cohort | 23-03-2020 | 26-04-2020 | 2019: same period | Jan-Feb 2020 VS Jan-Feb 2019 (adjacent non-pandemic period) | |
| 33 | Spain | Retrospective cohort | 04-03-2020 | 19-04-2020 | | 17-01-2020 to 03-03-2020 | |
| 34 | Greece | Retrospective cohort | 02-03-2020 | 12-04-2020 | 2019: same period | | |
| 35 | Italy | Retrospective cohort | 01-03-2020 | 31-03-2020 | 2019: same period | | |
| 36 | Germany | Retrospective cohort | 01-03-2020 | 30-04-2020 | 2019: same period | 01-01-2020 to 29-02-2020 | |
| 37 | Germany | Retrospective cohort | 21-03-2020 | 20-04-2020 | | | 2017-2019: same period |
| 38 | Austria | Retrospective cohort | 24-02-2020 | 05-04-2020 | | Comparison betweet the six weeks (1: calendar week 9-10, 2: week 11-12, 3: week 13-14) | |
| 39 | Multiple countries (b) | Survey | 01-04-2020 | 15-04-2020 | | | |
| 40 | Austria | Retrospective cohort | 01-01-2020 | 31-05-2020 | 2019: same period | | 2017-2018: same period |
| 41 | Italy | Retrospective cohort | 01-03-2020 | 31-03-2020 | 2019: same period | | |
| 42 | Multiple countries (c) | Systematic review/ Pooled analysis | | | 2019: same period | X | |

| | | | | | | | |
|---|---|---|---|---|---|---|---|
| 43 | Multiple countries (d) | Retrospective cohort | 01-03-2020 | 30-04-2020 | 2019: same period, (1 month: each investigator decided the relevant time period in his country) | | |
| 44 | US | Retrospective cohort | 04-03-2020 | 14-04-2020 | 2019: same period | 01-01-2020 to 03-03-2020 | |
| 45 | Italy | Retrospective cohort | 21-02-2020 | 10-04-2020 | | 3-1-2020 to 20-02-2020 | |
| 46 | Greece | Retrospective cohort | 15-03-2020 | 14-04-2020 | 2019: same period | | |
| 47 | Italy | Retrospective cohort + Literature review | 09-03-2020 | 09-04-2020 | 2019: same period | Previous months of 2020 | |
| 48 | US | Retrospective cohort | 01-01-2020 | 26-04-2020 | 2019: same period | | |
| 49 | United Kingdom | Retrospective cohort | 23-03-2020 | 19-04-2020 | | 01-01-2019 to 22-03-2020 (before Covid) | |
| 50 | Lithuania | Retrospective cohort | 11-03-2020 | 20-04-2020 | 2019: same period | | |
| 51 | US | Prospective cohort | 23-03-2020 | 15-04-2020 | | 01-01-2020 to 22-03-2020 | |
| 52 | Japan | Retrospective cohort | 07-04-2020 | 14-08-2020 | | | 01-01-2018 to 6-04-2020 |
| 53 | Switzerland | Retrospective cohort | 13-03-2020 | 30-04-2020 | 2019: same period | 07-01-2020 to 24-02-2020 | |
| 54 | Australia | Retrospective cohort | 16-03-2020 | 15-04-2020 | | | 2014-2019: same period |
| 55 | United Kingdom | Retrospective cohort | 19-02-2020 | 14-04-2020 | 2019: same period | | 2016-2018: same period |
| 56 | US | Survey | 15-03-2020 | 15-04-2020 | 2019: same period | | |
| 57 | Italy | Retrospective cohort | 20-02-2020 | 14-04-2020 | 2019: same period | | |
| 58 | United Kingdom | Retrospective cohort | 16-03-2020 | 16-05-2020 | | | 2017-2019: same period |

| | | | | | | | |
|---|---|---|---|---|---|---|---|
| 59 | Multiple countries (e) | Retrospective cohort | 01-03-2020 | 30-04-2020 | 2019: same period | | |
| 60 | Italy | Survey | 12-03-2020 | 19-03-2020 | 2019: same period | | |
| 61 | Canada | Retrospective cohort | mid-March 2020 | mid-May 2020 | 2019: same period | January to mid-March 2020 | |
| 62 | US | | 15-03-2020 | 25-04-2020 | | 01-01-2019 to 14-03-2020 | |
| 63 | France | Retrospective cohort | 17-02-2020 | 26-04-2020 | | | 2018-2019: same period |
| 64 | Israel | Prospective cohort | 20-03-2020 | 30-04-2020 | 2019: same period | | |
| 65 | United Kingdom | Retrospective cohort | 01-02-2020 | 30-04-2020 | | | 2017-2019 |
| 66 | United Kingdom | Retrospective cohort | 01-03-2020 | 30-04-2020 | 2019: same period | | |
| 67 | France | Prospective cohort | 09-03-2020 | 10-05-2020 | 2019: same period | | |
| 68 | Spain | Prospective cohort | 01-01-2020 | 10-05-2020 | 2019: same period | | |
| 69 | France | Prospective cohort | 26-02-2020 | 10-03-2020 | | | 2008-2017 |
| 70 | Spain | Retrospective cohort | 16-03-2020 | 14-04-2020 | 2019: 01-04 to 30-04 | | |
| 71 | Germany | Prospective cohort | 01-03-2020 | 31-03-2020 | | | 2017-2019: month of March |
| 72 | US | Retrospective cohort | 01-03-2020 | 30-04-2020 | 01-2019 to 02-2020 | | |
| 73 | Italy | Prospective cohort | 01-02-2020 | 31-05-2020 | 2019: same period | | 2017-2018: same period |
| 74 | Italy | Retrospective cohort | 01-03-2020 | 20-04-2020 | 2019: same period | | |
| 75 | Ireland | Prospective cohort | 01-03-2020 | 30-04-2020 | 2019: same period | Jan-Feb 2020 | |
| 76 | New Zealand | Prospective cohort | 30-12-2019 | 03-05-2020 | 2019: same period | | |
| 77 | Italy | Retrospective cohort | 01-03-2020 | 31-05-2020 | 2019: same period | | |
| 78 | Japan | Survey | 15-04-2020 | 30-04-2020 | | 3rd survey in 15-May | |
| 79 | United Kingdom | Retrospective cohort | 23-03-2020 | 30-04-2020 | | 01-01-2020 to 22-03-2020 | 2017-2019 |
| 80 | United Kingdom | Retrospective cohort | 01-01-2020 | 10-05-2020 | | | 2017-2019 |
| 81 | Spain | Survey | 01-01-2020 | 30-09-2020 | 2019: same period | | |

| | | | | | | | |
|---|---|---|---|---|---|---|---|
| 82 | Turkey | Retrospective cohort | 10-03-2020 | not clear | | before 13-01-2020 (not clear) | |
| 83 | Spain | Retrospective cohort | 15-03-2020 | 25-04-2020 | | | 2015-2019: same period |
| 84 | Multiple countries (f) | Survey | 01-03-2020 | 30-04-2020 | 2019: march | | |
| 85 | Multiple countries (g) | Survey | 15-06-2020 | 03-07-2020 | | | |
| 86 | United Kingdom | Survey | 15-06-2020 | 10-08-2020 | | | |
| 87 | US | Retrospective cohort | 15-03-2020 | 30-06-2020 | 2019: same period | | |
| 88 | France | Survey | 17-03-2020 | 27-03-2020 | | | |
| 89 | US | Retrospective cohort | 18-03-2020 | 02-06-2020 | 2019: same period | 01-01-2020 to 07-03-2020 | |

a) 17 Countries: Argentina, Mexico, Italy, US, Egypt, Spain, Perú, Austria, France, Saudi Arabia, Serbia, Poland, Turkey, Greece, Romania, Portugal, Uzbekistan, UK, Libya

b) Various countries - 49% Western Europe, 21% Eastern Europe, 13% Africa/Middle East, 11% Australasia and 6% Americas

c) 22 studies Europe, 8 USA, 3 China, 2 UK, 1 Canada, 1 Australia.

d) European centers - Poland, The Netherlands, Sweden, Portugal, Switzerland, Italy, Hungary, UK, Germany, Finland, Denmark, France, Spain, US

e) 77 European centers (5 European geographical areas were identified-1: Italy; 2: Iberian Peninsula (Spain and Portugal); 3: Central Europe (France,Germany,The Netherlands,Belgium,Czech Republic); 4:Balkan Peninsula (Romania,Slovenia,Greece,North Macedonia);5:North-East Europe(UK,Poland,Finland,Denmark,Russia).

f) 108 countries (Centers: Africa 42, Eastern Europe: 62, Far East: 99, Latin America: 201, MiddleEast/S.Asia: 60, US,Canada: 172, SoutheastAsia/Pacific:81, Western Europe: 192)

g) 23 countries (Excellence Center network of the European Society of Hypertension) - 20 European, 2Latin American,1Middle Eastern: (Italy,Greece,Spain,United Kingdom,Hungary,Austria, Brazil, France, Germany,Serbia, Sweden, Argentina,Armenia,Belgium,Bulgaria, Czech Republic, Estonia, Finland, Lebanon,Poland, Romania, Slovenia and Ukraine)

**Supplementary material 6 – Number of indicators and indicators' trends reported by each indicator, presented by country**

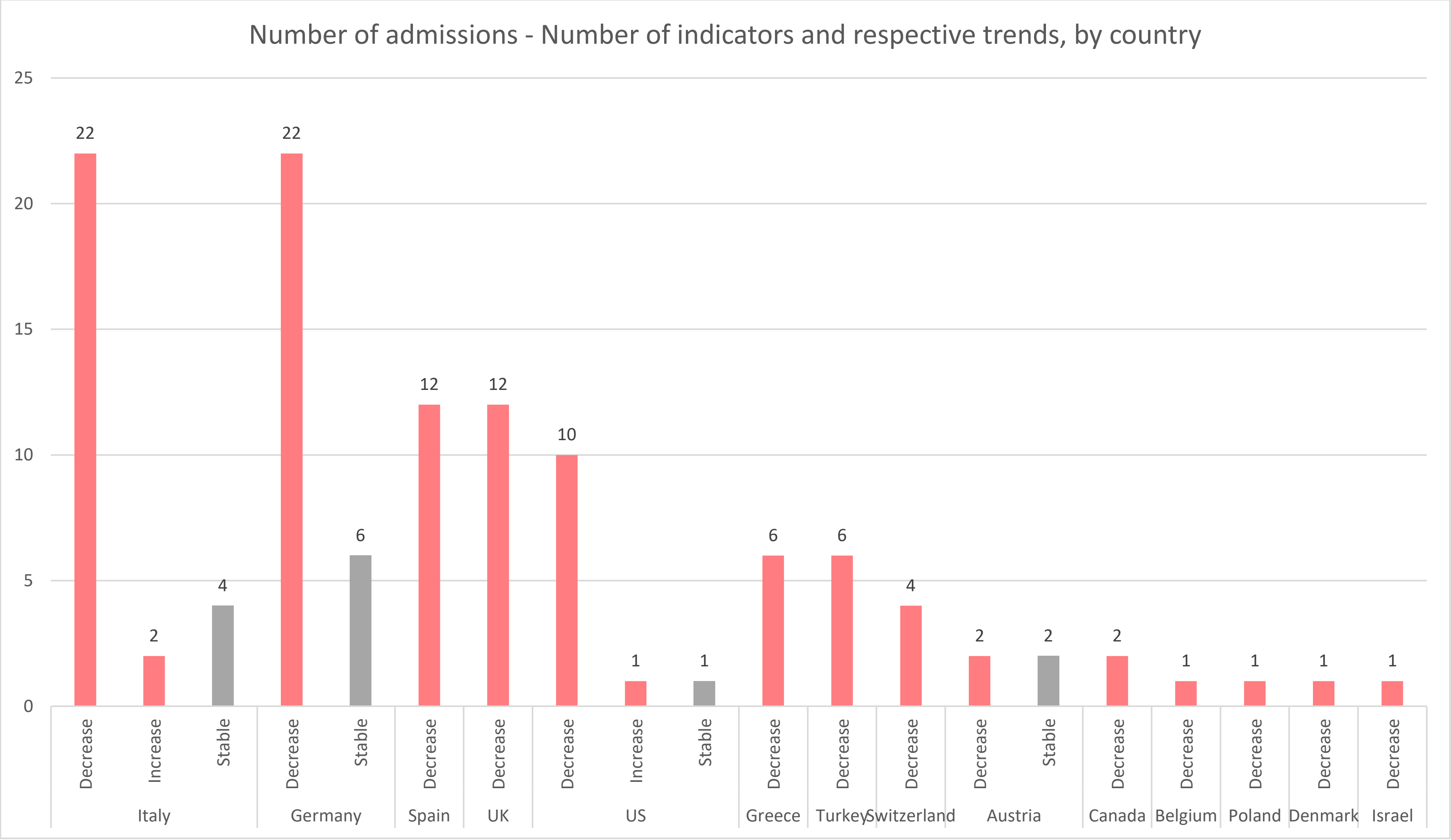

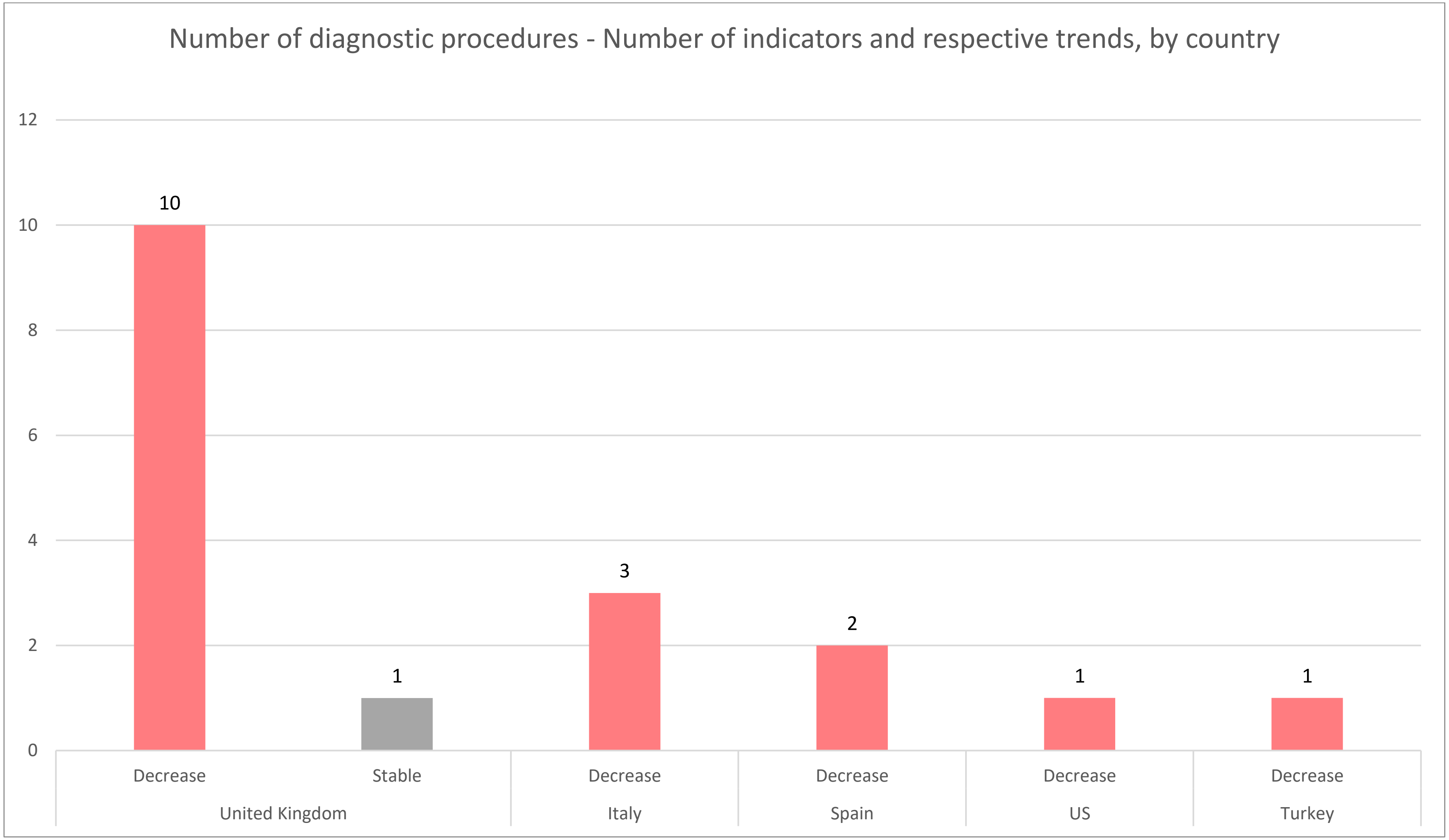
Number of diagnostic procedures - Number of indicators and respective trends, by country
12
10
8
6
4
2
0
10
1
3
2
1
1
Decrease
Stable
Decrease
Decrease
Decrease
Decrease
United Kingdom
Italy
Spain
US
Turkey

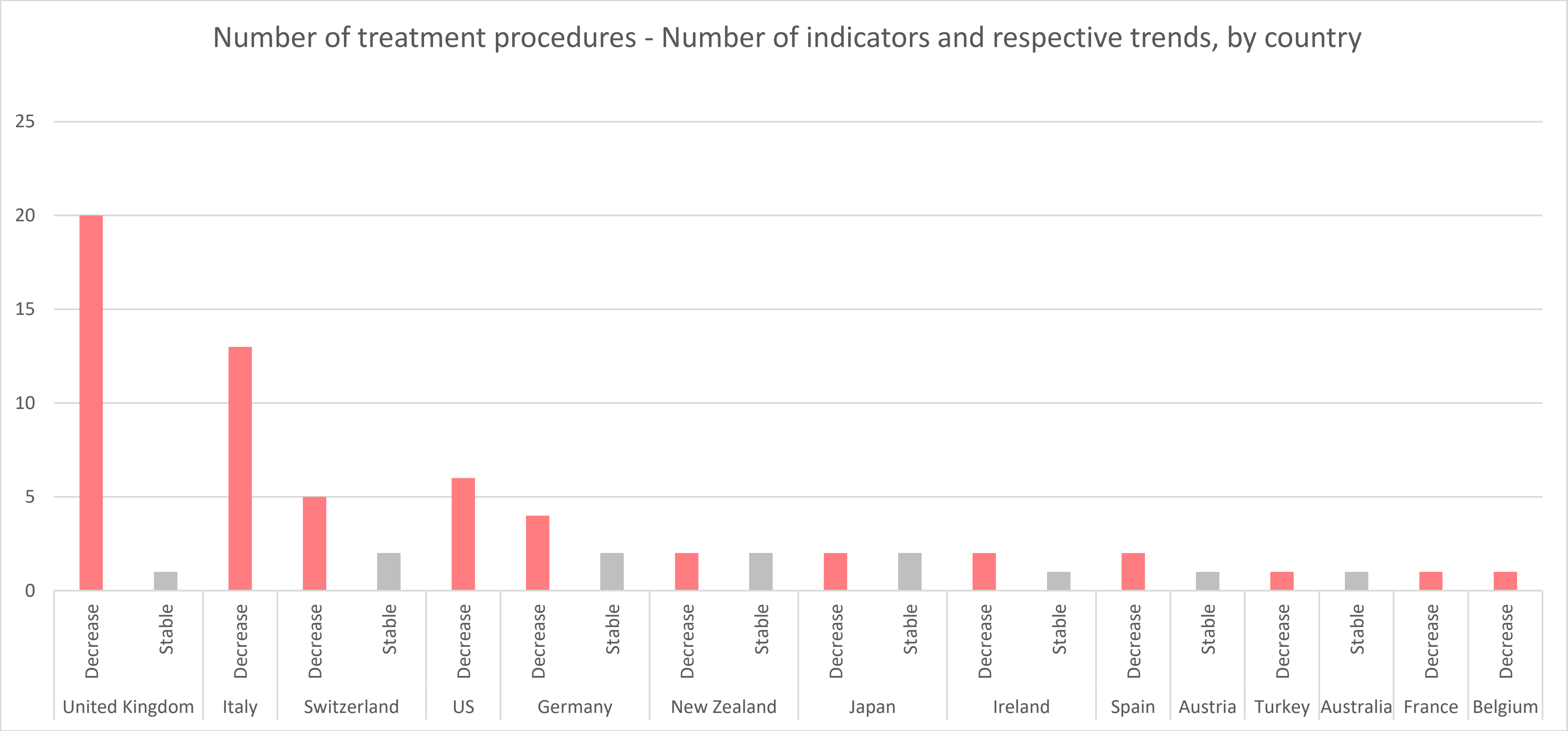

Number of treatment procedures - Number of indicators and respective trends, by country
25
20
15
10
5
0
Decrease
Stable
United Kingdom
Italy
Switzerland
US
Germany
New Zealand
Japan
Ireland
Spain
Austria
Turkey
Australia
France
Belgium

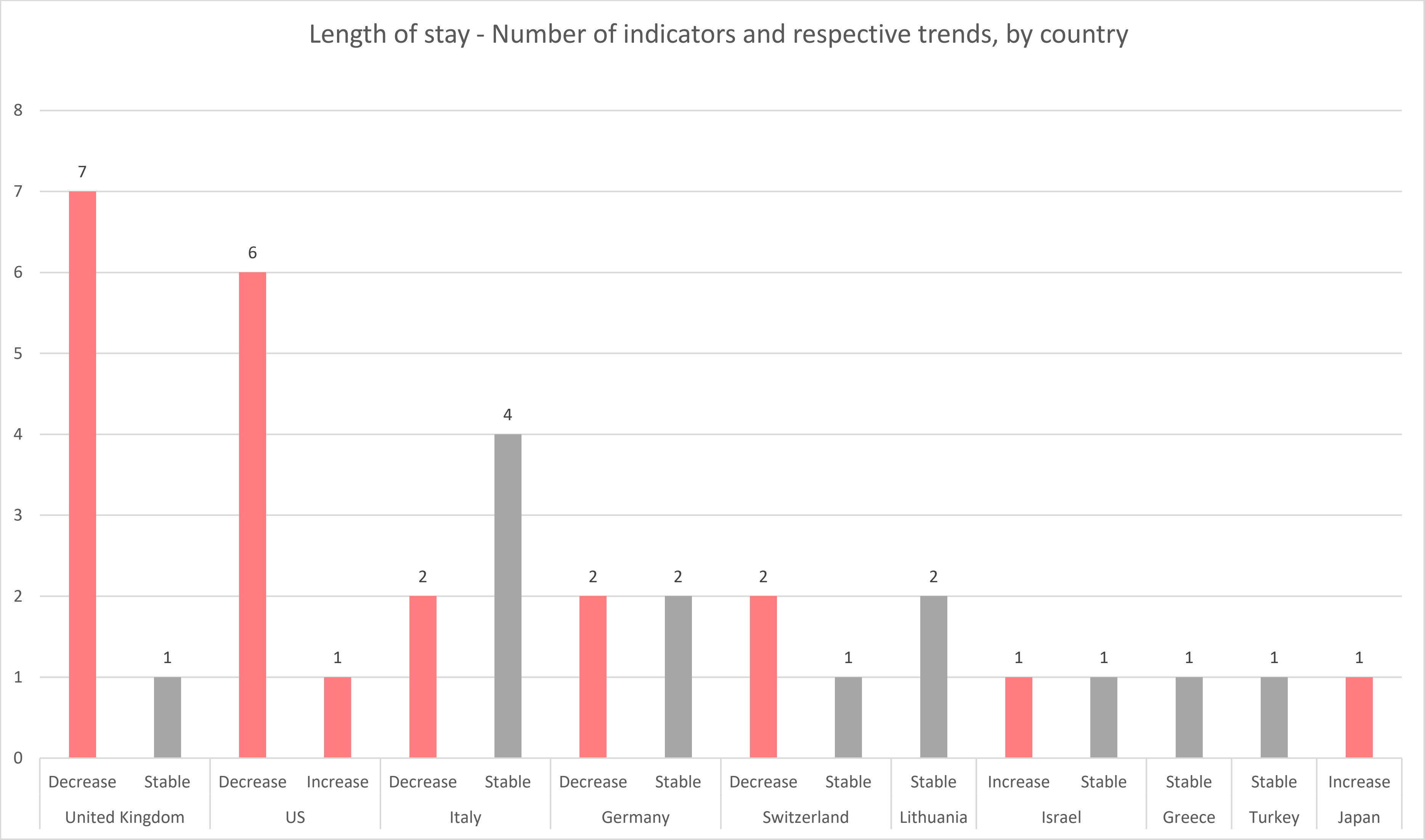
Length of stay - Number of indicators and respective trends, by country
0
1
2
3
4
5
6
7
8
7
1
6
1
2
4
2
2
2
1
2
1
1
1
1
1
Decrease
Stable
Decrease
Increase
Decrease
Stable
Decrease
Stable
Decrease
Stable
Stable
Increase
Stable
Stable
Stable
Increase
United Kingdom
US
Italy
Germany
Switzerland
Lithuania
Israel
Greece
Turkey
Japan

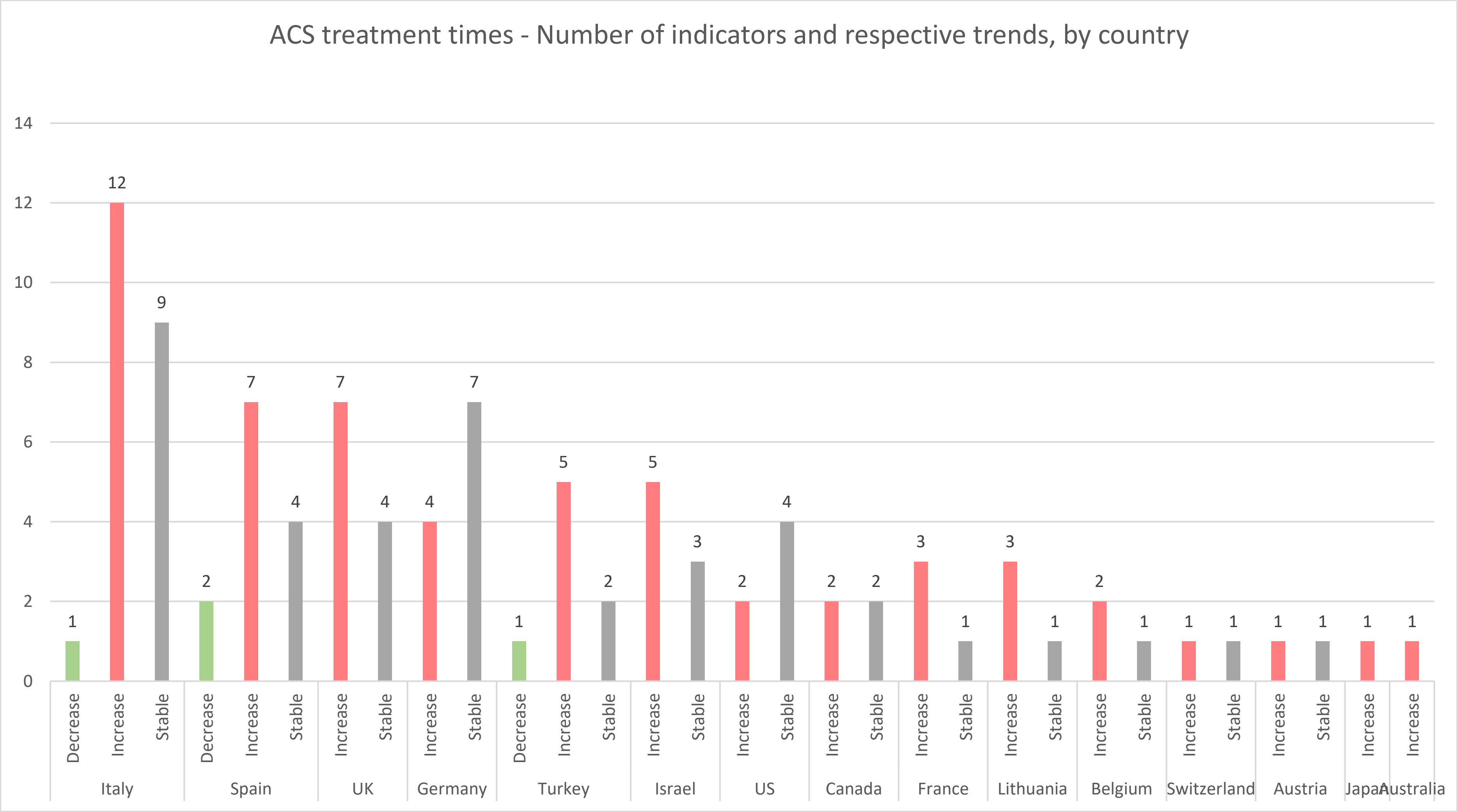
ACS treatment times - Number of indicators and respective trends, by country
0
2
4
6
8
10
12
14
1
12
9
2
7
4
7
4
4
7
1
5
2
5
3
2
4
2
2
3
1
3
1
2
1
1
1
1
1
1
1
Decrease
Increase
Stable
Italy
Spain
UK
Germany
Turkey
Israel
US
Canada
France
Lithuania
Belgium
Switzerland
Austria
Japan
Australia

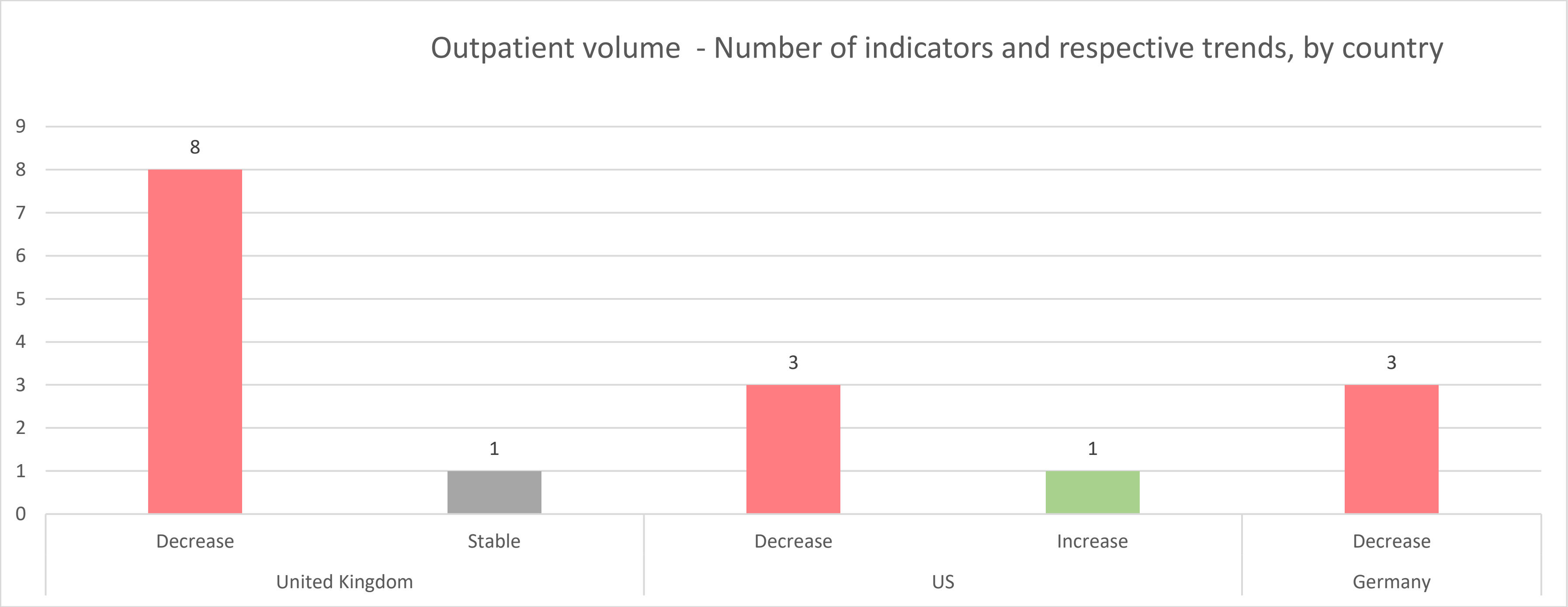
Outpatient volume - Number of indicators and respective trends, by country
9
8
7
6
5
4
3
2
1
0
8
1
3
1
3
Decrease
Stable
Decrease
Increase
Decrease
United Kingdom
US
Germany

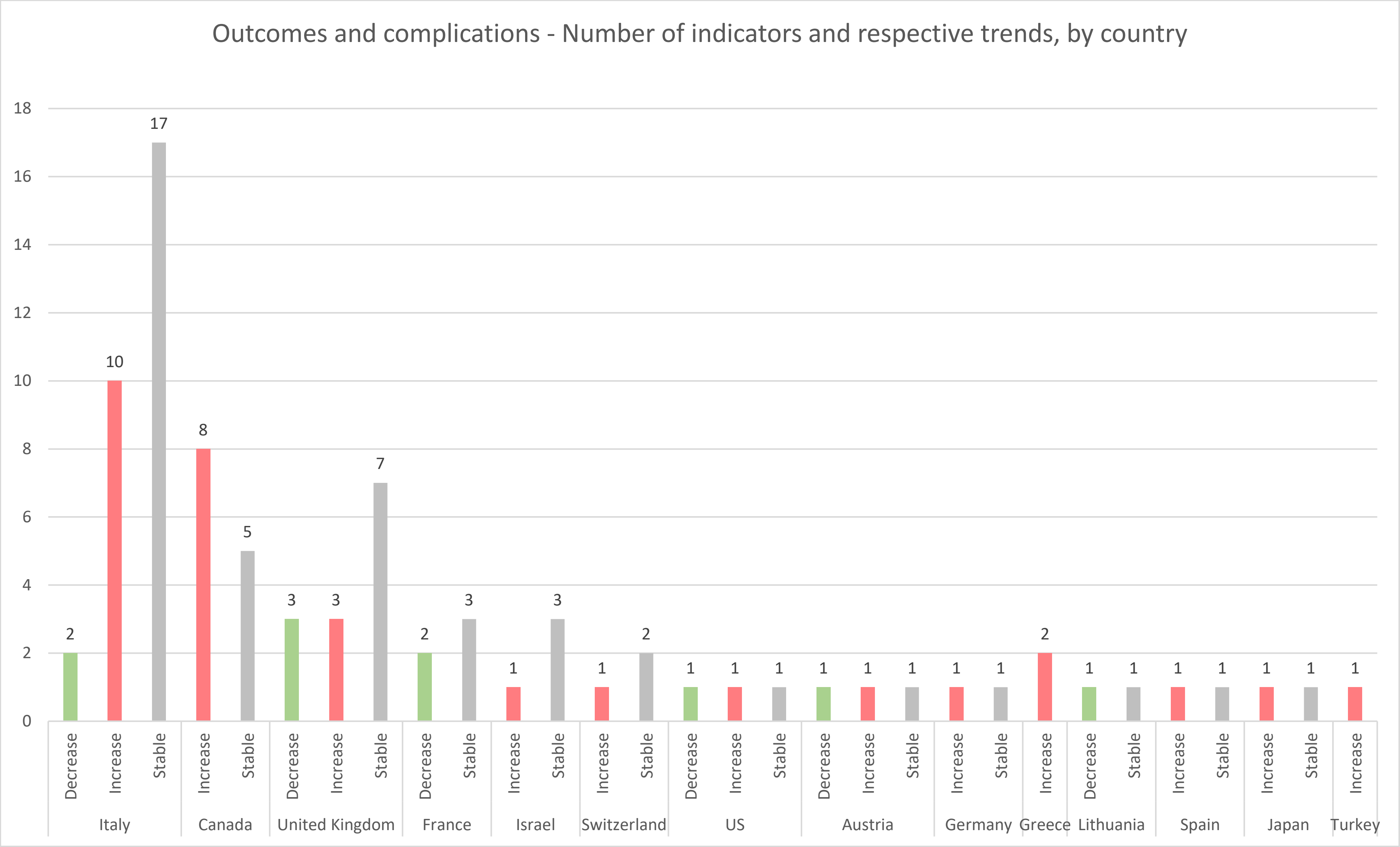

Outcomes and complications - Number of indicators and respective trends, by country
0
2
4
6
8
10
12
14
16
18
2
10
17
8
5
3
3
7
2
3
1
3
1
2
1
1
1
1
1
1
1
1
2
1
1
1
1
1
1
1
Decrease
Increase
Stable
Increase
Stable
Decrease
Increase
Stable
Decrease
Stable
Increase
Stable
Increase
Stable
Decrease
Increase
Stable
Decrease
Increase
Stable
Increase
Stable
Increase
Decrease
Stable
Increase
Stable
Increase
Stable
Increase
Italy
Canada
United Kingdom
France
Israel
Switzerland
US
Austria
Germany
Greece
Lithuania
Spain
Japan
Turkey

#### Supplementary material 7 – Additional references: studies included in the review, and from which indicators were extracted and collated

1. Alba A, Frankfurter C, Buchan T et al. REDUCED RATE OF HOSPITAL PRESENTATIONS FOR HEART FAILURE DURING THE COVID-19 PANDEMIC IN TORONTO, CANADA. *Can J Cardiol* 2020;36:S74.
https://www.ncbi.nlm.nih.gov/pmc/articles/PMC7529371/ 10.1016/j.cjca.2020.07.147

2. Araiza-Garaygordobil D, Montalto C, Martinez-Amezcua P et al. Impact of the COVID-19 Pandemic on Hospitalizations for Acute Coronary Syndromes: a Multinational Study. *QJM* 2021;10.1093/qjmed/hcab013

3. Ball S, Banerjee A, Berry C et al. Monitoring indirect impact of COVID-19 pandemic on services for cardiovascular diseases in the UK. *Heart* 2020;106:1890-1897.10.1136/heartjnl-2020-317870

4. Bhatt AS, Moscone A, McElrath EE et al. Fewer Hospitalizations for Acute Cardiovascular Conditions During the COVID-19 Pandemic. *J Am Coll Cardiol* 2020;76:280-288.10.1016/j.jacc.2020.05.038

5. Boeddinghaus J, Nestelberger T, Kaiser C et al. Effect of COVID-19 on acute treatment of ST-segment elevation and Non-ST-segment elevation acute coronary syndrome in northwestern Switzerland. *Int J Cardiol Heart Vasc* 2021;32:100686.10.1016/j.ijcha.2020.100686

6. Bollmann A, Hohenstein S, König S, Meier-Hellmann A, Kuhlen R, Hindricks G. In-hospital mortality in heart failure in Germany during the Covid-19 pandemic. *ESC Heart Failure* 2020;7:4416-4419.
https://onlinelibrary.wiley.com/doi/abs/10.1002/ehf2.13011 10.1002/ehf2.13011

7. Bromage DI, Cannatà A, Rind IA et al. The impact of COVID-19 on heart failure hospitalization and management: report from a Heart Failure Unit in London during the peak of the pandemic. *European Journal of Heart Failure* 2020;22:978-984.
https://onlinelibrary.wiley.com/doi/abs/10.1002/ejhf.1925 10.1002/ejhf.1925

36. Primessnig U, Pieske BM, Sherif M. Increased mortality and worse cardiac outcome of acute myocardial infarction during the early COVID-19 pandemic. *ESC Heart Failure* 2021;8:333-343.
https://onlinelibrary.wiley.com/doi/abs/10.1002/ehf2.13075 10.1002/ehf2.13075

37. Rattka M, Baumhardt M, Dreyhaupt J et al. 31 days of COVID-19-cardiac events during restriction of public life-a comparative study. *Clinical research in cardiology* 2020;109:1476-1482. https://www.ncbi.nlm.nih.gov/pubmed/32494921 10.1007/s00392-020-01681-2

38. Reinstadler SJ, Reindl M, Lechner I et al. Effect of the COVID-19 Pandemic on Treatment Delays in Patients with ST-Segment Elevation Myocardial Infarction. *J Clin Med* 2020;9 https://www.ncbi.nlm.nih.gov/pmc/articles/PMC7408681/ 10.3390/jcm9072183

39. Roffi M, Capodanno D, Windecker S, Baumbach A, Dudek D. Impact of the COVID-19 pandemic on interventional cardiology practice: results of the EAPCI survey. *EuroIntervention* 2020;16:247-250.10.4244/EIJ-D-20-00528

40. Schnaubelt S, Domanovits H, Niederdoeckl J et al. The Impact of the COVID-19 Pandemic on Incidences of Atrial Fibrillation and Electrical Cardioversion at a Tertiary Care Emergency Department: An Inter- and Intra-year Analysis. *Front Med (Lausanne)* 2020;7:595881.10.3389/fmed.2020.595881

41. Secco GG, Zocchi C, Parisi R et al. Decrease and Delay in Hospitalization for Acute Coronary Syndromes During the 2020 SARS-CoV-2 Pandemic. *Canadian Journal of Cardiology* 2020;36:1152-1155.
https://www.sciencedirect.com/science/article/pii/S0828282X2030502X 10.1016/j.cjca.2020.05.023

42. Singh S, Fong HK, Desai R, Zwinderman AH. Impact of COVID-19 on acute coronary syndrome-related hospitalizations: A pooled analysis. *Int J Cardiol Heart Vasc* 2021;32 https://www.ncbi.nlm.nih.gov/pmc/articles/PMC7836359/ 10.1016/j.ijcha.2021.100718

43. Sokolski M, Gajewski P, Zymliński R et al. Impact of Coronavirus Disease 2019 (COVID-19) Outbreak on Acute Admissions at the Emergency and Cardiology Departments Across Europe. *Am J Med* 2021;134:482-489.10.1016/j.amjmed.2020.08.043

44. Solomon MD, McNulty EJ, Rana JS et al. The Covid-19 Pandemic and the Incidence of Acute Myocardial Infarction. *N Engl J Med* 2020;383:691-693.10.1056/NEJMc2015630

45. Tomasoni D, Adamo M, Italia L et al. Impact of COVID-2019 outbreak on prevalence, clinical presentation and outcomes of ST-elevation myocardial infarction. *J Cardiovasc Med (Hagerstown)* 2020;21:874-881.10.2459/JCM.0000000000001098

46. Vassilikos VP, Pagourelias ED, Katsos K et al. Impact of social containment measures on cardiovascular admissions and sudden cardiac death rates during Coronavirus Disease (COVID-19) outbreak in Greece. *Hellenic J Cardiol* 2020;
https://www.ncbi.nlm.nih.gov/pmc/articles/PMC7491429/ 10.1016/j.hjc.2020.09.009

47. Vecchio S, Fileti L, Reggi A, Moschini C, Lorenzetti S, Rubboli A. Impact of the COVID-19 pandemic on admissions for acute coronary syndrome: review of the literature and single-center experience. *G Ital Cardiol (Rome)* 2020;21:502-508.10.1714/3386.33635

48. Vest AR, Upshaw JN, Dean K et al. The Decrease in Hospitalizations For Heart Failure During the Covid-19 Pandemic: A Community and Academic Hospital Comparison Study. *J Card Fail* 2020;26:S71.
https://www.ncbi.nlm.nih.gov/pmc/articles/PMC7527197/ 10.1016/j.cardfail.2020.09.207

49. Wu J, Mamas M, Rashid M et al. Patient response, treatments, and mortality for acute myocardial infarction during the COVID-19 pandemic. *European Heart Journal - Quality of Care and Clinical Outcomes* 2021;7:238-246.
https://doi.org/10.1093/ehjqcco/qcaa062 10.1093/ehjqcco/qcaa062

57. Barbieri L, Tumminello G, Lucreziotti S et al. Mortality in STEMI Patients During the COVID Era: Has the Pandemic Changed Our Clinical Practice? *Cardiovasc Revasc Med* 2021;22:120-121.
https://www.ncbi.nlm.nih.gov/pmc/articles/PMC7488593/ 10.1016/j.carrev.2020.09.014

63. Hauguel-Moreau M, Pillière R, Prati G et al. Impact of Coronavirus Disease 2019 outbreak on acute coronary syndrome admissions: four weeks to reverse the trend. *J Thromb Thrombolysis* 2020;:1-2.
https://www.ncbi.nlm.nih.gov/pmc/articles/PMC7323878/ 10.1007/s11239-020-02201-9
